# Altered model-based prediction error signaling in the lateral orbitofrontal cortex in patients with obsessive-compulsive disorder: An fMRI study

**DOI:** 10.1101/2024.07.17.24310561

**Authors:** Pritha Sen, Kathrin Koch, Benita Schmitz-Koep, Deniz Gürsel, Franziska Knolle

**Author notes:** corresponding author: Dr. Franziska Knolle, Department of Diagnostic and Interventional Neuroradiology, School of Medicine and Health, Klinikum rechts der Isar, Technical University Munich, 81675 Munich, Germany.

## Abstract

**Background:** Compared to healthy individuals, patients with obsessive-compulsive disorder (OCD) are found to rely more on model-free decision-making strategies which may underlie symptom expression. It is, however, unclear whether these behavioral differences are represented in neural alterations of model-free and model-based decision-making when tested simultaneously.

**Methods:** We investigated the neural signatures of 22 OCD patients and 22 matched controls who completed a two-step Markov decision-making task during functional MRI scanning. We used hierarchical Bayesian modelling and Bayesian statistics to examine model-based and model-free decision-making behaviors. Parametric regressors were employed for model-free and model-based reward prediction errors to inform neural reward presentation, which we analyzed using a Bayesian Multilevel Modeling (BML) approach. Associations between significant activations and symptoms as well as cognitive scores were explored using Bayesian linear regression.

**Results:** While controls received significantly more rewards and were significantly less stochastic compared to patients, both groups similarly relied on model-free decision-making strategies. Importantly, our group comparison of neural reward prediction error responses showed greater activation for model-based reward prediction error in the lateral orbitofrontal cortex (lateral OFC) in OCD patients compared to controls, but no differences for model-free reward prediction error processing. Increased lateral OFC activity was associated with lower obsessive symptoms and better cognitive functioning.

**Conclusion:** These findings support the notion that OCD is associated with an altered goal-directed system, which may be expressed through increased activation in the lateral OFC activity underlying goal-directed behavior. Importantly, the hyperactivity observed in this region was linked to reduced obsessive symptoms and improved cognitive functioning, potentially indicating compensatory mechanisms and highlighting the lateral OFC as a potential target for future interventions.

## 1. Introduction

One of the key impairments of obsessive-compulsive disorder (OCD) is the inability to make effective and advantageous decisions (Robbins et al., 2019; Sachdev and Malhi, 2005). Decision-making allows successful interaction with the environment through planning, monitoring, feedback integration and feedback-based updating. This process can be categorized into two distinct systems – model-free and model-based decision-making, which operate in parallel to choose the most suitable action (Daw et al., 2005; Gläscher et al., 2010; Grosskurth et al., 2019).

Model-free decision-making is reinforcement-learning based, and therefore automatic, guided by past successful choices, and inflexible to changes (Balleine and O’Doherty, 2010; Huang et al., 2020; Voon et al., 2015a). Model-free decision-making may be described as habit learning, which requires direct responses to environmental stimuli without following a goal or explicit value. Model-based decisions, on the other hand, require an understanding of action-outcome contingencies, and with this an understanding of a given task. They are, therefore, deliberate, flexible, and adaptive, enabling individuals to optimize their decisions by anticipating the consequences of their actions (Balleine and O’Doherty, 2010; Daw et al., 2011; Gillan and Robbins, 2014), which relates to goal-directed behavior. Both model-free and model-based decision-making utilize reinforcement learning principles, relying on prediction errors to guide behavior, aiming for optimal outcomes. However, model-free decision-making relies on past rewards to guide behavior, producing automatic and habitual responses to environmental stimuli, and is inflexible to changes (Balleine and O’Doherty, 2010; Huang et al., 2020; Voon et al., 2015a). It may be described as habit learning, which requires direct responses to environmental stimuli without following a goal or explicit value. Model-based decisions, on the other hand, require an understanding of action-outcome contingencies and the structure of a given task. Model-based learning aligns with goal-directed behavior, as it represents causal relationships between actions and their consequences, which is particularly relevant in scenarios requiring contingency awareness. They may be described as a form of instrumental conditioning, where outcomes are induced by explicit actions. Distinct neural systems represent these goal-directed and model-based actions (Balleine and O’Doherty, 2010; Robbins et al., 2024). A recent meta-analysis (Huang et al., 2020) revealed overlapping but distinct regions, showing that while the goal-directed system has been linked to activity in the ventral striatum, the medial prefrontal cortex (mPFC) and the orbitofrontal cortex (OFC), the habit system has been associated with activity in ventral striatum as well, but also in the globus pallidus and caudate head. Within the OFC, the lateral and medial OFC specifically play important and distinct roles in goal-directed decision-making (Frank and Claus, 2006; Kim, 2021; Noonan et al., 2017). The medial OFC is involved in sustaining consistent stimulus-outcome associations, while the lateral OFC tracks dynamic outcomes to facilitate adaptive response adjustments (Wallis, 2007; Windmann et al., 2006).

Individuals with OCD are usually characterized by a relatively high inflexibility and tendency towards sticking to habits and routines. Thus, it does not come as a surprise that they often exhibit an imbalance in these two systems, favoring rigid and repetitive decisions (Gillan et al., 2011a; Gillan and Robbins, 2014; Robbins et al., 2019), which lead to inflexibility and resistance to change (Gillan and Robbins, 2014; Morein-Zamir et al., 2013; Murray et al., 2019; Wheaton et al., 2019), potentially due to an overreliance on model-free decision-making. Additionally, compulsions in OCD are also shown to arise from this core difficulty of regulating actions and thoughts in a goal-directed manner (Gillan, 2017; Nestadt et al., 2016; Pushkarskaya et al., 2015; Robbins, 2022). Neuroimaging studies looking at model-based and model-free decision-making separately, suggest that this shift towards habitual decisions in OCD is associated with alterations in the cortico-striato-thalamo-cortical (CSTC) circuits. Activity within these loops, involving – amongst others – OFC, anterior cingulate cortex (ACC) and caudate nucleus, which are responsible for governing habits and automated behaviors (Burton et al., 2015; Huang et al., 2020; Miller et al., 2018; Simmler and Ozawa, 2019), have been proposed to become hyperactive or unresponsive to inhibitory signals in OCD, potentially establishing the neuronal basis for the incapacity to inhibit obsessive or compulsive thoughts or behaviors (Burguière et al., 2015; Graybiel and Rauch, 2000). Additionally, reduced grey matter volume and decreased activity in the medial prefrontal cortex (mPFC), a region responsible for constructing, updating and maintaining an internal model of the environment to guide goal-directed behavior (Ahmari and Rauch, 2022; Seow et al., 2021; Szalisznyó and Silverstein, 2021), has been linked to over-reliance on habitual control in OCD patients (Ahmari and Rauch, 2022; Ruan et al., 2023). Furthermore, disruption of activity in various regions of the basal ganglia, such as, subthalamic nucleus, nucleus accumbens, has been found to be associated with impaired behavioral inhibition, resulting in heightened impulsive choices (Christakou et al., 2004; Jahanshahi et al., 2015).

Few studies investigate model-based and model-free decision-making simultaneously in OCD (Kim et al., 2024; Voon et al., 2015a). The two-step Markov decision-making task (Daw et al., 2011) distinguishes between model-free and model-based learning behaviorally, and when combined with fMRI, revealed that the striatal reward prediction error (RPE) signal reflected both model-free and model-based decisions in healthy individuals (Daw et al., 2011). In a behavioral set-up of this two-step task, Voon, Derbyshire and colleagues (Voon et al., 2015b) found that individuals with different compulsive behaviors (binge eating disorders, methamphetamine addiction, and OCD patients), resorted to more model-free decisions. A follow-up behavioral study comparing the influence of reward and loss outcomes on decisions in this task demonstrated that, following rewarded trials, OCD patients leaned more towards model-free decisions, while the reverse was observed in loss trials (Voon et al., 2015a). Notably, patients displayed a heightened tendency to maintain the same first-level choice in consecutive trials, irrespective of win or loss, compared to healthy controls. Furthermore, the severity of compulsion scores correlated with habitual learning in rewarded trials whereas obsession scores correlated with increased choice switching following loss trials. In a very recent study, Kim and colleagues (Kim et al., 2024), for the first time, explored neurocomputational alteration during model-based and model-free decision-making in OCD patients using the two-step task and applying a computational model which was designed to account for an uncertainty-based arbitration process of model-based and model-free strategies. They found, first, that patients showed hypoactivity in the inferior frontal gyrus when tracking the reliability of goal-directed behavior; and second, that, patients exhibited weaker right ipsilateral ventrolateral prefronto-putamen coupling to reach specific goals, which was correlated with more severe compulsivity. More evidence, however, is needed to understand the neural mechanisms of altered model-based and model-free decision-making and their relevance for clinical symptoms in OCD.

Therefore, the aim of this study was to explore the neural mechanisms of model-based and model-free decision-making in OCD patients using the two-step task with fMRI, incorporating hierarchical Bayesian modelling and Bayesian Multilevel Modeling (BML) which performs Bayesian analyses based on pre-specified regions of interest. All other statistical analyses were also performed using Bayesian statistical methods, following a timely debate of misinterpretations of p-values and the replication crisis (Anderson, 2020). Bayesian statistical approaches incorporate prior information into the analysis allowing for regularization of models and preventing overfitting, which increases the precision of the findings, making them more generalizable and reliable (Edwards et al., 1963; Van De Schoot et al., 2017). Based on the literature, our regions of interest included the caudate, putamen, nucleus accumbens, mPFC, OFC and ACC for both model-free and model-based RPE processing. Timeseries of the model-free and model-based decision-making were used to explore which strategy was reflected in each region and whether this differed across groups. We hypothesized that the OCD patients would exhibit predominantly model-free behavior with caudate, putamen, nucleus accumbens and ACC activations reflecting more model-free RPE, while the healthy controls would display a mixture of model-free and model-based behavior, with the mPFC, OFC and the nucleus accumbens reflecting more model-based RPE. Furthermore, we expected that neural alterations would link to symptom severity. We also expected that neural alterations in model-based prediction-errors would be associated with stronger compulsivity.

## 2. Methods

### 2.1. Participants

Twenty-five adults with OCD and 23 healthy controls were recruited for this study out of which one healthy control and three OCD patients had to be excluded from the final analysis due to corrupted imaging data. The final data for the analysis, therefore, consisted of 22 OCD patients and 22 healthy controls. We conducted a Bayes Factor Design Analysis to retrospectively assess the adequacy of our sample size. Using a medium effect size (δ = 0.5) and a decision boundary of Bayes Factor ≥ 3 (moderate evidence), we observed that a minimum sample size of 22 participants per group is needed to achieve robust evidence. Demographic information, clinical and cognitive scores are presented in Table 1. Patients met Diagnostic and Statistical Manual of Mental Disorders (DSM)-IV criteria for OCD (First et al., 2002). Exclusion criteria for both groups were a history of severe head injuries, seizures, neurological diseases, schizophrenia, autism, substance and alcohol abuse/dependency, mental retardation, severe medical conditions, and pregnancy. 16 patients were under medication while the other six were medication-naïve or had stopped medication at least one week before scanning. The study was approved by the Technical University of Munich ethics committee, and all participants gave informed consent after receiving a complete description of the study.

**Table 1.** Demographic characteristics, clinical characteristics and cognitive functioning scores of healthy controls and patients with obsessive-compulsive disorder.

|  | OCD | Healthy<br>Controls | Group Differences |
| --- | --- | --- | --- |
| Demographic characteristics |  |  |  |
| Age (SD) | 33.4 (11.8) | 32.2 (8.33) | B=-0.66, 95% CI [-6.5, 5.05] |
| Gender | 17F, 5M | 13F, 9M | B=-5.7, 95% CI [-15.22, -2.3] |
| Clinical scores |  |  |  |
| Years since onset | 14.32 (9.41) |  |  |
| Age of onset | 19.04 (10.06) |  |  |
| Y-BOCS Obsessions | 11.2 (4.16) |  |  |
| Y-BOCS Compulsions | 10 (4.23) |  |  |
| Y-BOCS total | 21.2 (7.04) |  |  |
| OCI-R | 26.9 (9.47) |  |  |
| HAMD | 12.5 (4.6) |  |  |
| Cognitive scores |  |  |  |
| TMT-A (s) | 29 (13.2) | 22 (6.10) | B=-5.69, 95% CI [-11.79, 0.34] |
| TMT-B (s) | 66.5 (31.5) | 56.1 (18.9) | B=-9.48, 95% CI [-26.4, 7] |
| TMT (B-A) (s) | 37.5 (23) | 34.1 (17.4) | B=-3.18, 95% CI [-15.45, 9.65] |
| Note: Data are presented as mean (SD) except for Gender. Abbreviations: M=Male; F=Female; SD=Standard Deviation; TMT=Trail-making test; Y-BOCS=Yale-Brown Obsessive-Compulsive Scale; OCI-R=Obsessive-Compulsive Inventory – Revised; HAMD=Hamilton rating scale for depression; B = Bayesian estimate; CI not containing zero indicates substantial effect. |  |  |  |

### 2.2. Task description

All the participants performed the two-step task by Daw and colleagues (Daw et al., 2011) inside an fMRI scanner, which is a reinforcement learning task to measure the relative degree of model-free against model-based decision-making. The task comprised of 150 trials divided into three runs. Each trial in the task consisted of two stages. In both the stages, the subjects were presented with two fractal images as stimuli to choose from using an MR-compatible button box. In stage 1, two images (A and B, refer to Figure 1) were displayed amongst which they were required to select one. Depending on their choice in stage 1, a set of two images were presented to them on stage 2. The two possible sets of images in stage 2 were set ‘a’ and set ‘b’, consisting of two fractal images in each set. On common transitions, image A when chosen on stage 1, led to set ‘a’ on stage 2, and image B normally led to set ‘b’, with probability of 70%. However, in 30% of the trials, an uncommon transition occurred in which image A when chosen on stage 1, led to set ‘b’ on stage 2, and image B led to ‘set a’. Common and uncommon transitions were randomly distributed across all the trials. In stage 2, the subject was required to choose one image from the set, after which the subject either received a reward or no reward before starting with the next trial. The reward probability changed slowly and independently based on Gaussian random walks within the range of 0.25 to 0.75 (Figure 1). Based on receiving or not receiving a reward in stage 2, the participants were expected to develop a strategy that maximizes the chances of obtaining a reward.

**Figure 1.**
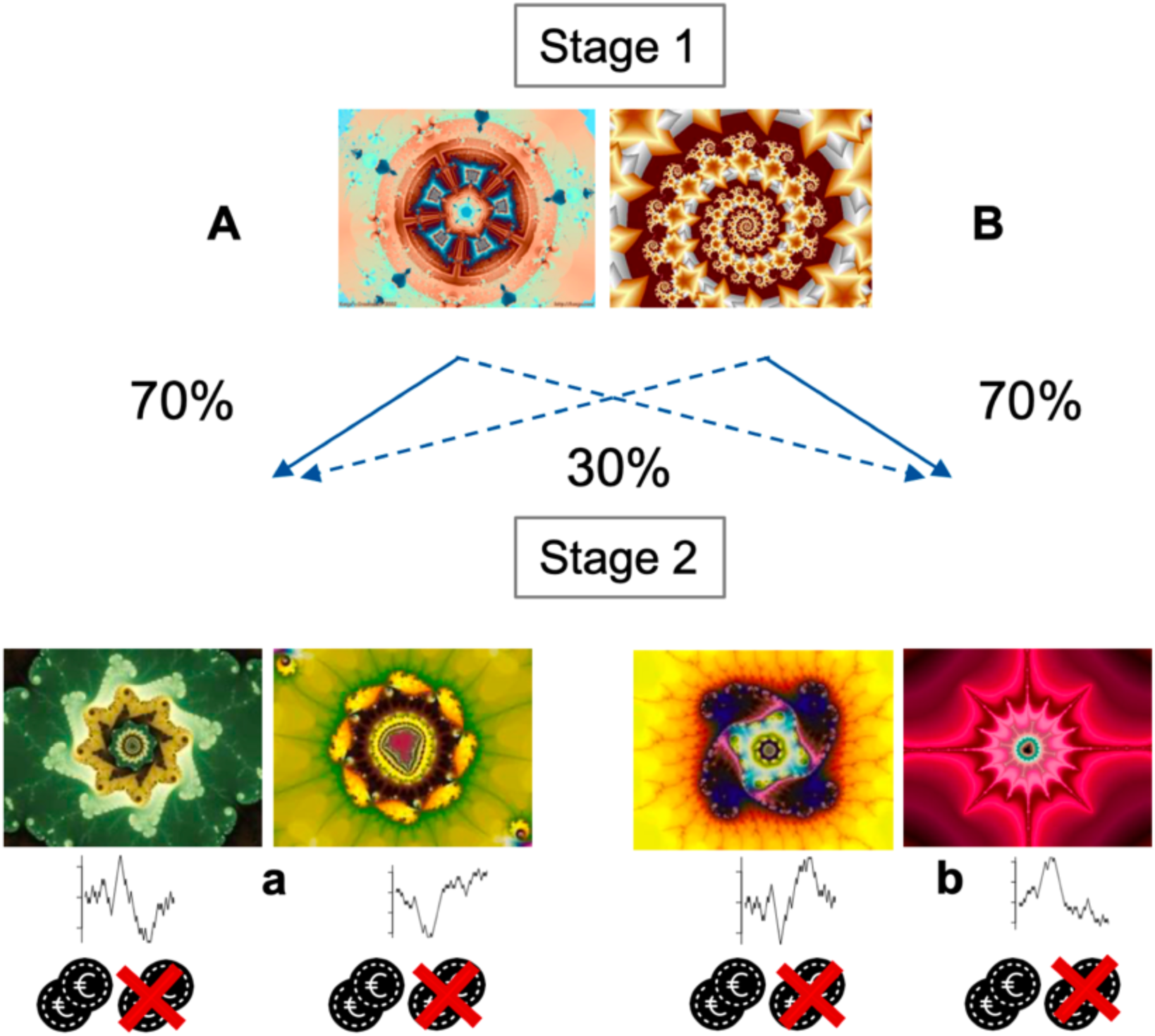
Two-step Markov decision task. On each trial, a first stage choice between two images (here, A or B) leads probabilistically to a second stage choice (here, images in set ‘a’ or images in set ‘b’). Importantly, each first-stage stimulus is more strongly (70% vs 30%) associated with a particular second-stage state throughout the experiment, imposing a task structure that can be exploited in a model-based choice. All stimuli in stage 2 are associated with a probabilistic reward changing slowly and independently based on Gaussian random walks, forcing subjects to continuously learn and explore the second stage choices.

To make a model-based decision, the subject must use knowledge of transition probabilities between the states to infer the likelihood that a particular action will lead to a reward. In model-free behavior, on the other hand, the transition behavior between the states is not considered and the current value of each action is updated on the basis of whether actions were rewarded in the preceding trial. Therefore, trials after rare transitions are used to distinguish between model-based and model-free actions.

The first-stage stimuli were presented for a max. of 5s. The choice of the first-stage was presented for 1.5s. The second-stage stimuli were also presented for a max. of 5s. Feedback presentation after the second-stage was 2s. The inter-trial intervals were randomized between 2-6 s.

### 2.3. Behavioral Analysis

#### 2.3.1. Task performance

Reaction times (in seconds) for stage 1 and stage 2 responses were calculated for both groups and compared using robust Bayesian ANOVA. Additionally, to measure performance in the task, reward percentages for both groups were calculated as the ratio of number of rewarded trials to the total number of trials per subject and compared using robust Bayesian ANOVA, followed by a Bayes factors (BF_10_) analysis, quantifying the more likely hypothesis based on the observed data. Evidence for alternative hypothesis over null hypothesis was identified if BF_10_ > 1 (BF_10_ [1-3]: anecdotal evidence; BF_10_ [3-10]: moderate evidence; BF_10_ [10-30]: strong evidence; BF_10_ [30-100]: very strong evidence; BF_10_ > 100: extreme evidence).

#### 2.3.2. Stay probability

To identify differences between patients and controls towards a decision bias, we assessed the probability for staying with the same stage 1 choice in the subsequent trial which is deterministic of model-free and model-based choices. See supplementary material Section 1.3. for details. Stay probabilities for ideal model-free and model-based performances were also simulated along and compared visually with actual stay probability in the task.

We used Bayesian regression analysis, assuming normal distribution, to investigate whether the stay probability varied based on participant groups (healthy controls, OCD patients), previous trial transition type (common or uncommon), previous trial outcome (rewarded or unrewarded), as well as their interactions. As we had multiple observations per person, we incorporated a group level intercept to account for the resulting dependency in the data. A substantial evidence (refer to Section 2.5 for details on identifying a substantial evidence) in the interaction between transition type and outcome would reflect model-based decisions, and lack of a substantial evidence of interaction would indicate model-free decisions that is purely guided by reward without considering the transition structure. The equation for the model that we fit is as follows:

stay probability ∼ previous trial reward · previous trial transition type · participants + (1|Subject)

#### 2.3.3. Computational modelling of task behavior

Hierarchical Bayesian modelling was employed on the behavioral data (Ahn et al., 2017) to understand the underlying mechanisms of decision-making in OCD patients compared to healthy controls and inform the imaging analysis. We fitted four models, each with 4 chains, 1000 burn-in samples and 3000 samples, after which model selection for further analysis was determined based on model-convergence and lowest leave one out information criterion (LOOIC) values. The LOOIC evaluates model fit by iteratively excluding individual observations, i.e. single trials or data points within each subject to check the predictive accuracy of the model without those observations. The 6-parameter model was found to be the winning model and was used for further analysis. See supplementary material Section 2.1., 2.2., and 2.3. for description of models, fitting procedures, model architectures and model comparisons. Supplementary Table 1 provides descriptions of the parameters generated by the four models; Supplementary Table 2 provides LOOIC values for all the fitted models; Supplementary Figures 2-4 present caterpillar plots showing model convergence for all models fitted across healthy controls, OCD patients, and all subjects, respectively; and Supplementary Figure 5 presents box plots showing the distribution of individual parameter estimates for the two groups from the winning model. To quantify how much variance in behavior the hierarchical Bayesian model explains, we also performed a Bayesian regression analysis to assess how well the parameters from the winning 6-parameter model explained the stay probability behavior (See supplementary material 2.5. for more details). Supplementary Figure 9 presents posterior predictive checks supporting the validity of the regression model, comparing observed stay probabilities with model replicated values.

The 6-parameter model estimates the following parameters:

**Learning rate (‘alpha 1 and alpha 2’):** showing the efficiency of learning over the trials in stage 1 and stage 2, takes values from 0 to 1. The higher the score, the better the subjects understand and perform in the task.

**Inverse-temperature/choice stochasticity (‘beta1 and beta2’):** referring to the proportions of stochastic/random choices made during the task in stage 1 and stage 2, with beta=0 for completely random responding and beta=∞ for stochastically choosing the highest value option.

**Perseverance (‘pi’):** Perseverance determines how strongly the subject(s) stick to their decisions. Higher the score on the perseverance scale, lower the chance of switching to a different image.

**Model-weights (‘w’):** referring to the degree of model-based influence on choices, takes values from 0 to 1, ‘0’ indicating more model-free decisions and ‘1’ being more model-based.

Trial-by-trial model-regressors were extracted from the winning computational model for both model-free and model-based prediction errors to be used as parametric modulators in the imaging analysis (Daw et al., 2011). However, prediction error signals in the brain often reflect overlapping contributions from both model-free and model-based systems, making it challenging to interpret the respective contributions of each process. Without accounting for this shared variance, the neural signals regressed on the simple model-based regressor might also reflect model-free influences. See Supplementary Figure 6 for model-free regressors across trials and Supplementary Figure 7 for model-based regressors across trials, both displaying strikingly similar patterns. To isolate the true model-based signal, we calculated a difference regressor by subtracting model-free prediction errors from the model-based RPEs, as explained by Daw et al., (2011). This approach reduces multicollinearity by minimizing shared variance between the two regressors, and isolates unique contributions from the model-based system (see Supplementary Figure 8). By accounting for this shared variance, the difference regressor enables a more precise identification of neural activity associated with purely model-based processes. Please see supplementary material for more details on the model regressors (Section 2.4.).

To investigate group differences for the model parameters, we employed Bayesian robust one-way ANOVAs with each parameter as the dependent variable and group as the predictor variable, followed by a Bayes factors (BF_10_) analysis.

### 2.4. Imaging Analysis

#### 2.4.1 Image acquisition

A 3T Phillips scanner with a 32-channel head coil was used to acquire MRI data. T * weighted images were obtained using multiband echo-planar imaging with the following parameters: repetition time (TR): 2700 ms, echo time (TE): 30 ms, flip angle (FA): 90°, field of view (FOV): 192 × 141 × 192 mm^2^, matrix size: 96 × 94, number of slices: 64, number of volumes: 500, slice thickness: 2mm; gap: 0.6 mm; multiband factor = 2. High-resolution anatomical T1-weighted images were acquired using a magnetization-prepared rapid acquisition gradient echo (MPRAGE) sequence (TR: 11.18 ms; TE: 4.6 ms, FA: 8°, FOV = 250 × 250 × 88 mm, matrix size = 256 × 128, scan duration = 45.5 s, slice thickness = 2 mm). In order to take T1 equilibration effects into account, the first five volumes of every run were automatically discarded.

#### 2.4.2 Image pre-processing

Subsequent pre-processing of images and data analysis was done using the Statistical Parametric Mapping software, SPM12 (Ashburner et al., 2021). The functional data for each participant was slice-time corrected and realigned to each run’s mean functional image using a 6 degree-of-freedom rigid body spatial transformation after which the resulting images were co-registered to the participant’s structural image. The structural data was then co-registered to the functional data and segmented into gray and white-matter probability maps. These segmented images were used to calculate spatial normalization parameters to the ICBM152 template, which were subsequently applied to the functional data. The data was also resampled to 2 mm × 2 mm × 2 mm as a part of spatial normalization. The structural image was normalized to standard MNI space, and the warps were applied to the functional images. The functional images were then spatially smoothed using a 5 mm Gaussian kernel.

#### 2.4.3. fMRI Analysis

After preprocessing, subject-specific design matrices were defined using general linear modelling as implemented in SPM12 (Ashburner et al., 2021). The fMRI analysis was centered around analyzing the time-series of model-free and model-based RPEs. Four conditions were entered to calculate the average BOLD responses across all trials at each of the four time points in the task (i.e., onset first stage images, button-press first stage, onset second stage images, and button-press with reward presentation on the second stage). The computationally derived reward prediction error signals (mf_RPE and mfb_RPE) were entered as parametric regressors (please refer to supplementary material Section 2.4. for more details) in the fourth condition, i.e. the button-press second stage with reward presentation. Motion parameters were included as nuisance regressors and slow signal drifts with a period longer than 128 seconds were removed using a high pass filter. For the second-level models, the contrasts of interest defined were ‘model-free reward prediction error’ and ‘model-based reward prediction error’. The per-subject estimate of the model-based effect (*‘w’* obtained from the computational model – Section 2.3.3.) was also included as a second-level covariate for the model-based regressor to test the correspondence between behavioral and neural estimates of the model-based effect. These contrasts were taken to a second-level one-sample t-test across all controls and patients.

The second level analysis was performed on specific regions of interest that have been implicated in model-free and model-based RL in past studies – caudate, putamen, nucleus accumbens, medial prefrontal cortex (mPFC), anterior cingulate cortex (ACC), medial orbitofrontal cortex (medial OFC) and lateral orbitofrontal cortex (lateral OFC) (Figure 3A). Anatomical masks of the caudate, putamen and nucleus accumbens were derived from the Harvard-Oxford Subcortical Atlas and created and combined using FSL (Jenkinson et al., 2012); anatomic masks for the ACC was created combining Brodmann areas (BA) 24 and 32, anatomical mask for the medial OFC included BA 11 and that for lateral OFC included BA 47. Daw and colleagues (2011) identified the mPFC coordinate for model-free and model-based decisions to be x=-4, y=60, z=14, which corresponds to the BA 10, also recognized as part of the mPFC in the literature (Grossmann, 2013; Peng et al., 2018; Tavares, 2020). Therefore, BA 10 served as the mPFC mask in our study. Masks defined using BA were constructed as implemented in the WFU PickAtlas Toolbox in SPM. Parameter estimates for these ROIs were extracted from the second level contrast images using MarsBaR in SPM and entered into the Region-Based Analysis Program through BML (Chen et al., 2019), in AFNI (Cox, 1996).

#### 2.4.4. BML Analysis

As the BML is a recent neuroimaging technique, a concise overview is presented here (for a deeper understanding, refer to Chen and colleagues (Chen et al., 2019)). BML involves estimating the likelihood of a hypothesis based on observed data, represented as a probability density function termed the posterior distribution (abbreviated as P+). This distribution is derived by combining empirical data with a model and prior expectations. BML offers the advantage of consolidating data across subjects and Regions of Interest (ROIs), facilitating the integration of shared information among these regions. The more the posterior distribution deviates from 0, the stronger the evidence for an effect. The evidence for effects with a probability (P+) of <.10 or >.90 as categorized as weak, <.05 or >.95 as moderate, and <.025 or >.975 as strong. A probability (P+) of >.10 or <.90 is considered no evidence. Effects with a P+ of <.10 or >.90 are discussed in the main text, while comprehensive effects are presented in the probability distribution plots.

#### 2.4.5. Associations between neural responses and clinical and cognitive scores

To examine the relationships between clinical and cognitive scores, and brain activations in patients that were significantly different from those in controls, we conducted Bayesian linear regressions for each of the symptom (i.e., Y-BOCS obsession and compulsion, OCI-R scores, HAMD) and cognitive scores (i.e., TMT-A, TMT-B, TMT-BA) as the outcome variable and significant brain activations as predictors covarying for age of illness onset.

Y-BOCS obsessions measures the severity of obsessive thoughts, and Y-BOCS compulsions assesses the severity of compulsive behaviors (Goodman et al., 1989). Higher scores indicate more frequent or intense obsessions and compulsions, respectively. The OCI-R is a self-report measure that evaluates overall OCD symptom severity across various domains, including checking, washing, ordering, obsessing, and hoarding, with higher scores indicating more severe OCD symptoms (Foa et al., 2002). HAMD assesses the severity of depressive symptoms, where higher scores representing more severe depressive symptoms (Williams, 1988).

TMT-A and TMT-B are measures of processing speed (Sánchez-Cubillo et al., 2009), and executive functioning and cognitive flexibility (Arbuthnott and Frank, 2000), respectively. In TMT-A, subjects are required to connect numbers, while in TMT-B they are requested to alternate between numbers and letters in an ascending order as quickly as possible. The difference score, TMT-(B-A), is an index of executive functioning independent of processing speed (Corrigan and Hinkeldey, 1987; Sánchez-Cubillo et al., 2009). Higher scores on the TMT indicate higher degree of cognitive impairment.

The response variables were assumed to follow a Gaussian distribution for the residuals, and priors for the regression coefficients were specified using a normal distribution with a mean of 0 and standard deviation of 10. The posterior distributions were sampled using the No-U-Turn Sampler with 4 chains, each containing 2000 iterations, including a 1000-iteration warmup phase.

### 2.5. Substantial evidence for an effect

In Bayesian regression and Bayesian ANOVA analysis, an effect was identified by inspecting the credibility intervals (CI). If the CI did not contain 0, a substantial evidence for an effect was detected.

### 2.6. General statistical implementations

All statistical analyses were performed using R Statistical Software (version 4.3.1) (R Core Team, 2021); hierarchical Bayesian modelling was performed using hBayesDM (Ahn et al., 2017); Bayesian linear regression and ANOVA analysis was carried out using the brms package (version 2.20.4) (Bürkner, 2017); imaging analysis was performed using Statistical Parametric Mapping software, SPM12 (Ashburner et al., 2021); masks for region of interest analysis was made using FMRIB Software Library (FSL) (version 6.0.5.2) (Jenkinson et al., 2012); and BML analysis for the regions of interest was performed using the Region Based Analysis Program (Chen et al., 2019) in the Analysis of Functional NeuroImages (AFNI) Software (version 23.2.12) (Cox, 1996). Please see supplementary material Section 1.1. for more details.

## 3. Results

### 3.1. Task performance differences across groups

We did not observe any group differences in the reaction times (in seconds) for stage 1 (Healthy controls – Mean: 0.87, SD: 0.17, OCD patients – Mean: 0.88, SD: 0.14) and stage 2 (Healthy controls – Mean: 1.10, SD: 0.23, OCD patients – Mean: 1.16, SD: 0.32) responses.

The robust Bayesian ANOVA revealed a group difference in reward percentage for healthy controls (Mean: 60.3, SD: 3.43) and OCD patients (Mean: 54.6, SD: 3.5) (B=5.61, EE=1, 95% CI [3.64, 7.57]), with Bayes Factor analysis revealing extreme evidence (BF_10_=4779.379), indicating that controls performed better than OCD patients.

Additionally, to illustrate that participants remained engaged with the task throughout, we plotted cumulative reward accumulation across trials for each group (Supplementary Figure 1). We observed a steady increase in cumulative reward across trials in both groups, including in the later blocks of the task, suggesting sustained task engagement. While cumulative reward is not a direct measure of learning in the two-step task, since rewards are determined by independent Gaussian random walks, it serves as a general indicator that participants were interacting with the task meaningfully. Please see supplementary material Section 1.2. for details.

### 3.2. Stay probability differences across groups

Results of the Bayesian regression analysis revealed no main effect for group, but a main effect of reward (rewarded vs. unrewarded: B=0.14, EE=0.03, 95% CI [0.07, 0.21]), indicating that all participants stayed on the same first level choice in the next trial if the previous trial was rewarded, suggesting a model-free behavior. We found no evidence for interaction effects indicating that the subjects followed a model-free decision-making approach. Stay probabilities for ideal model-free and model-based behavior are displayed in Figure 2a and actual stay probabilities in the current task are displayed in Figure 2b.

**Figure 2.**
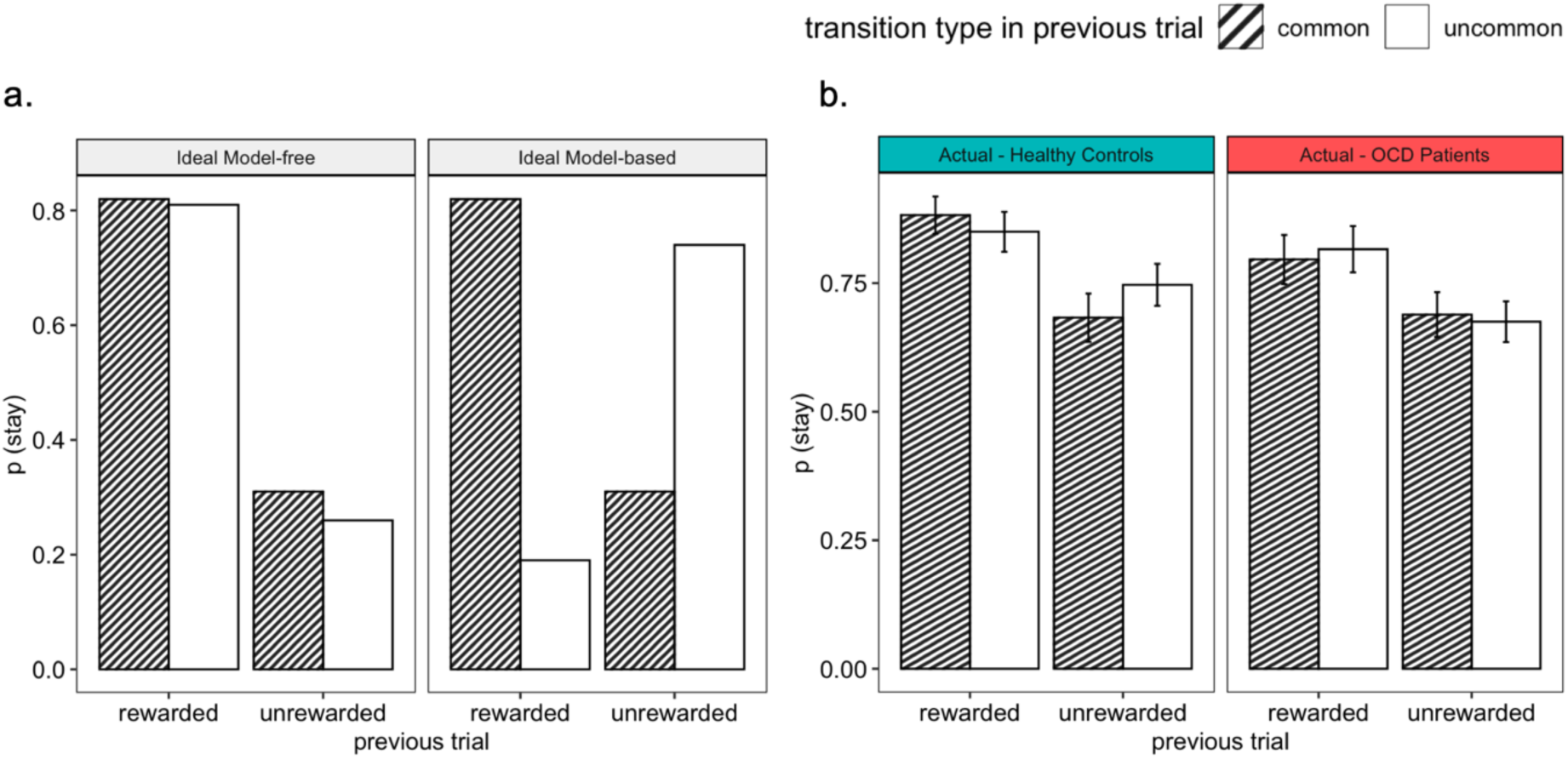
Ideal and actual stay-switch behaviors. **a.** Ideal Model-free and Ideal Model-based decision-making behavior: Model-free RL predicts that a first-stage choice yielding a reward is likely to be repeated on the upcoming trial, regardless of a common or an uncommon transition, Model-based RL predicts that an uncommon transition should affect the value of the next first stage option, leading to a predicted interaction between reward and transition probability. **b.** Actual stay proportions, averaged across subjects, displaying a characteristic of using both model-free and model-based decision-making strategies.

### 3.3. Computational modelling parameters differences across groups

We did not observe group differences in any of the computational modelling parameters described in Section 2.3.3., except ‘beta 1’. The robust Bayesian ANOVA revealed group differences in stochasticity at stage 1 ‘beta 1’ (healthy controls vs. OCD patients: B=1.57, EE=0.7, 95% CI [0.21, 2.94]). The Bayes Factor analysis for stochasticity at stage 1 ‘beta 1’ scores indicated anecdotal evidence (BF_10_=2.65), suggesting that patients were more random in their choices compared to controls. None of the other parameters revealed substantial effects.

### 3.4. Differences in the neural signature of model-free and model-based prediction errors

In the ROI BML analysis, group comparisons between healthy controls and OCD patients in the model-based RPE signal revealed that the OCD patients had a higher activation in the lateral OFC compared to healthy controls (weak evidence; P+=0.09, 95% CI [-3.42, 0.68]). We did not find any group differences in the model-free RPE signal in patients and controls. Parameter estimates for group comparisons in model-based and model-free RPE signal are displayed in Figure 3B.

**Figure 3.**
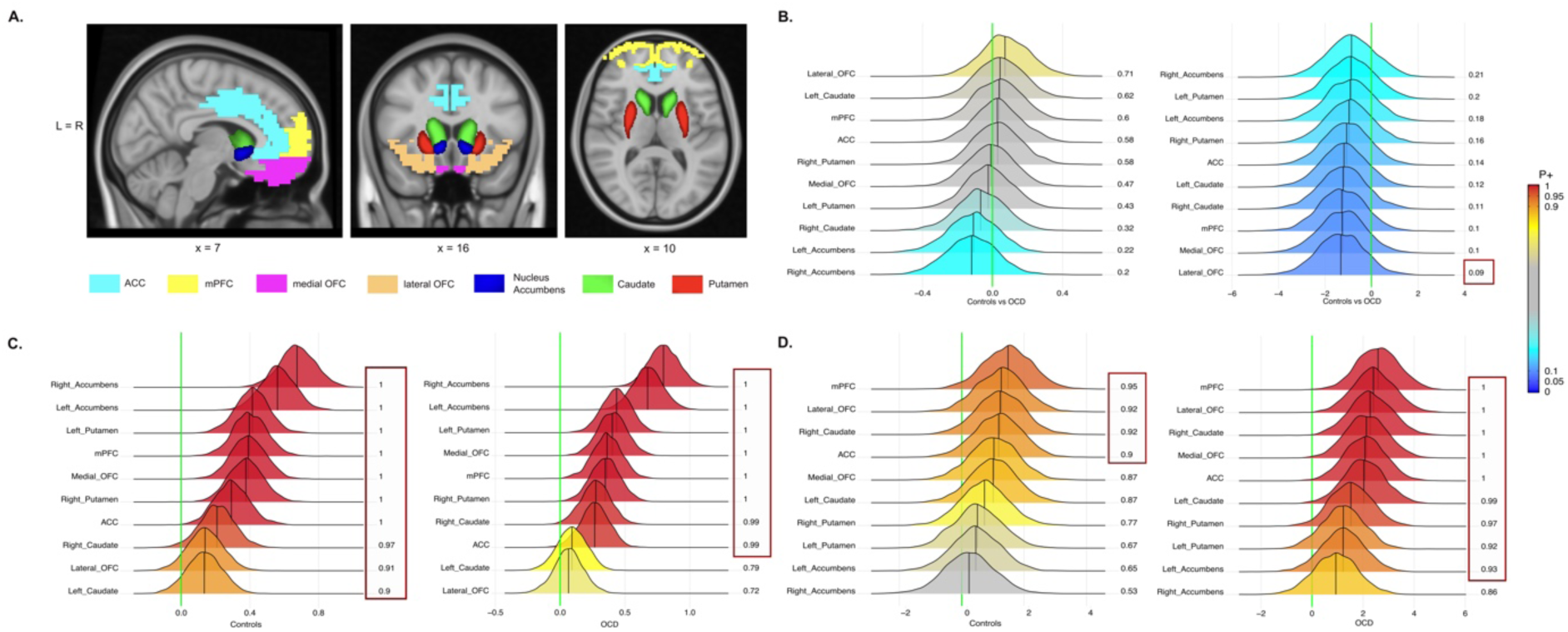
ROI analysis. **A. Regions included in the ROI analysis.** ROIs for model-free and model-based RPE; Masks were derived from the Harvard-Oxford Subcortical Atlas using FSL. **B. ROI analysis showing posterior distributions for OCD patients compared with healthy controls (left:** Model-free RPE; **right:** Model-based RPE). Values >.90 represent higher activation in healthy controls, and <.10 represent lower activation in the healthy controls. **C. ROI analysis showing posterior distributions within each group for model-free RPE** (**left:** Healthy controls; **right:** OCD patients). **D. ROI analysis showing posterior distributions within each group for model-based RPE** (**left:** Healthy controls; **right:** OCD patients). Note: Posterior distributions: strong evidence: (P+ > 0.975 or <0.025), moderate evidence (P+ > 0.95 or <0.05), weak evidence (P+ >0.90 or <0.10) of an effect. Boxes outlined in red highlight regions with substantial effects in each of these plots.

Within group effects for model-based RPE revealed activations in the mPFC (weak evidence; P+=0.95, 95% CI [-0.27, 2.83]), lateral OFC (weak evidence; P+=0.92, 95% CI [-0.43, 2.59]), right caudate (weak evidence; P+=0.92, 95% CI [-0.46, 2.59]), and ACC (weak evidence; P+=0.902, 95% CI [-0.55, 2.47]) in healthy controls; and activations in the mPFC (strong evidence; P+=1, 95% CI [0.97, 4.2]), lateral (strong evidence; P+=1, 95% CI [0.82, 4.08]) and medial (strong evidence; P+=1, 95% CI [0.57, 3.77]) OFC, right (strong evidence; P+=1, 95% CI [0.67, 3.96]) and left (strong evidence; P+=0.99, 95% CI [0.42, 3.64]) caudate, ACC (strong evidence; P+=1, 95% CI [0.51, 3.63]), right (moderate evidence; P+=0.97, 95% CI [-0.06, 3.13]) and left (weak evidence; P+=0.92, 95% CI [-0.44, 2.81]) putamen, and left accumbens (weak evidence; P+=0.93, 95% CI [-0.48, 2.84]) in the OCD patients.

Within group effects of model-free RPE revealed activations in the mPFC (strong evidence; Controls: P+=1, 95% CI [0.2, 0.6], OCD: P+=1, 95% CI [0.12, 0.58]), right accumbens (strong evidence; Controls: P+=1, 95% CI [0.46, 0.89], OCD: P+=1, 95% CI [0.57, 1.03]), left accumbens (strong evidence; Controls: P+=1, 95% CI [0.36, 0.76], OCD: P+=1, 95% CI [0.44, 0.91]), left putamen (strong evidence; Controls: P+=1, 95% CI [0.21, 0.61], OCD: P+=1, 95% CI [0.21, 0.65]), right putamen (strong evidence; Controls: P+=1, 95% CI [0.18, 0.58], OCD: P+=1, 95% CI [0.12, 0.57]), medial OFC (strong evidence; Controls: P+=1, 95% CI [0.19, 0.59], OCD: P+=1, 95% CI [0.17, 0.63]) ACC (strong evidence; Controls: P+=1, 95% CI [0.1, 0.51], OCD: P+=0.99, 95% CI [0.04, 0.48]) and right caudate (weak evidence in Controls; P+=0.97, 95% CI [-0.01, 0.42], strong evidence in OCD; P+=0.99, 95% CI [-0.05, 0.5]) for both healthy controls and OCD patients. In healthy controls, we also observed activations in the lateral OFC (weak evidence; P+=0.91, 95% CI [-0.06, 0.34]) and left caudate (weak evidence; P+=0.902, 95% CI [-0.07, 0.33]). Parameter estimates for group comparisons in model-based and model-free RPE signal for healthy controls and OCD patients are displayed in Figures 3C and 3D respectively.

### 3.5. Bayesian Linear Regressions results

We examined the relationships between model-based RPE signal in the lateral OFC, OCD symptoms and cognitive scores in OCD patients (Table 2). The Bayesian regression revealed that lateral OFC activity significantly and negatively predicted Y-BOCS obsessions (B=-0.98, EE=0.14, 95% CI [-1.85, –0.09]), TMT-A (B=-0.99, EE=0.45, 95% CI [-1.90, –0.11]) and TMT-B (B=-2.36, EE=1.05, 95% CI [-4.4, –0.30]). This indicates that higher model-based RPE signal in the lateral OFC is linked to reduced obsessive symptoms, faster processing speed, executive functioning and worse cognitive flexibility. All these Bayesian regressions were controlled for age of onset. No other regression analyses revealed significant results.

**Table 2.**
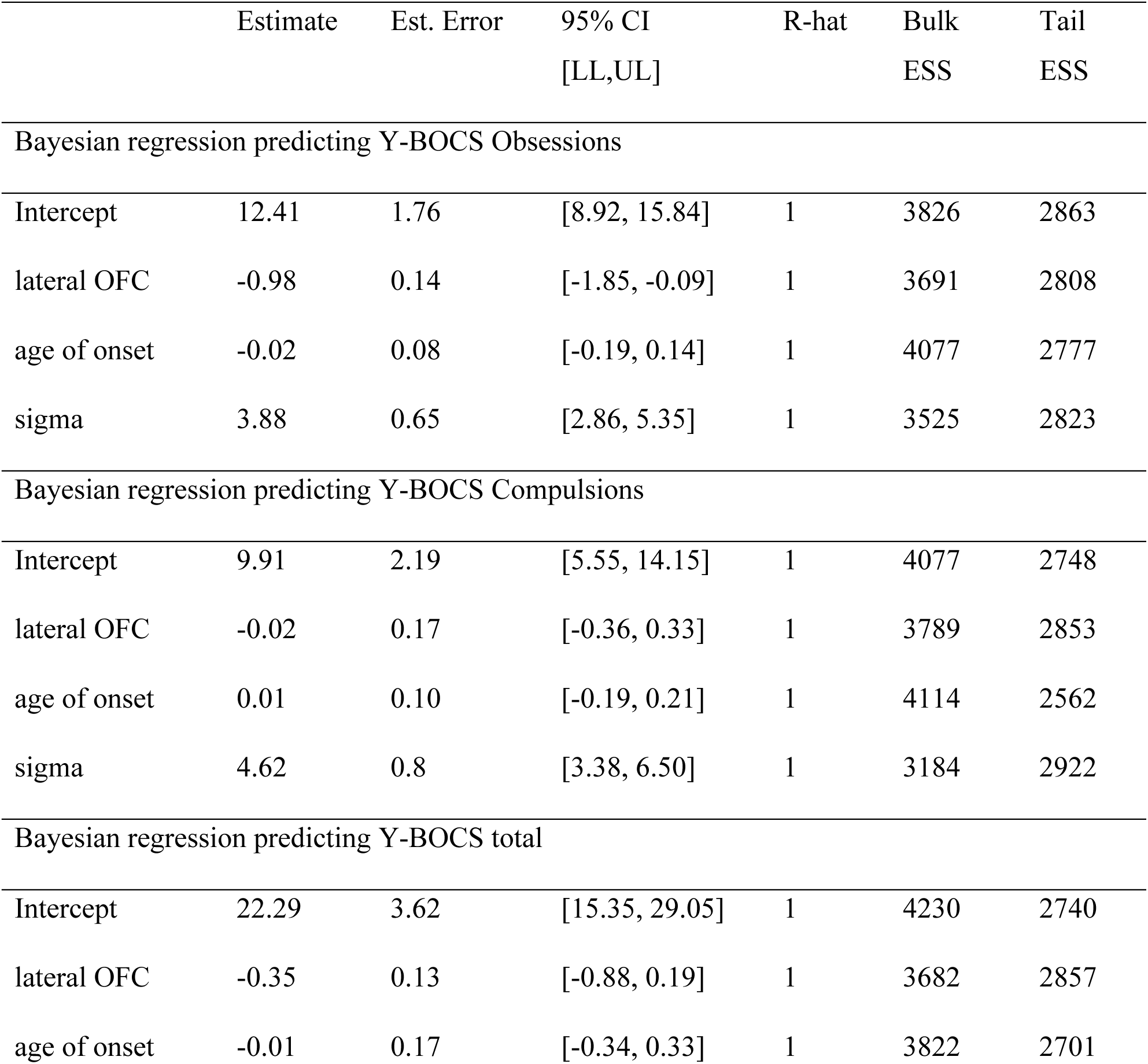
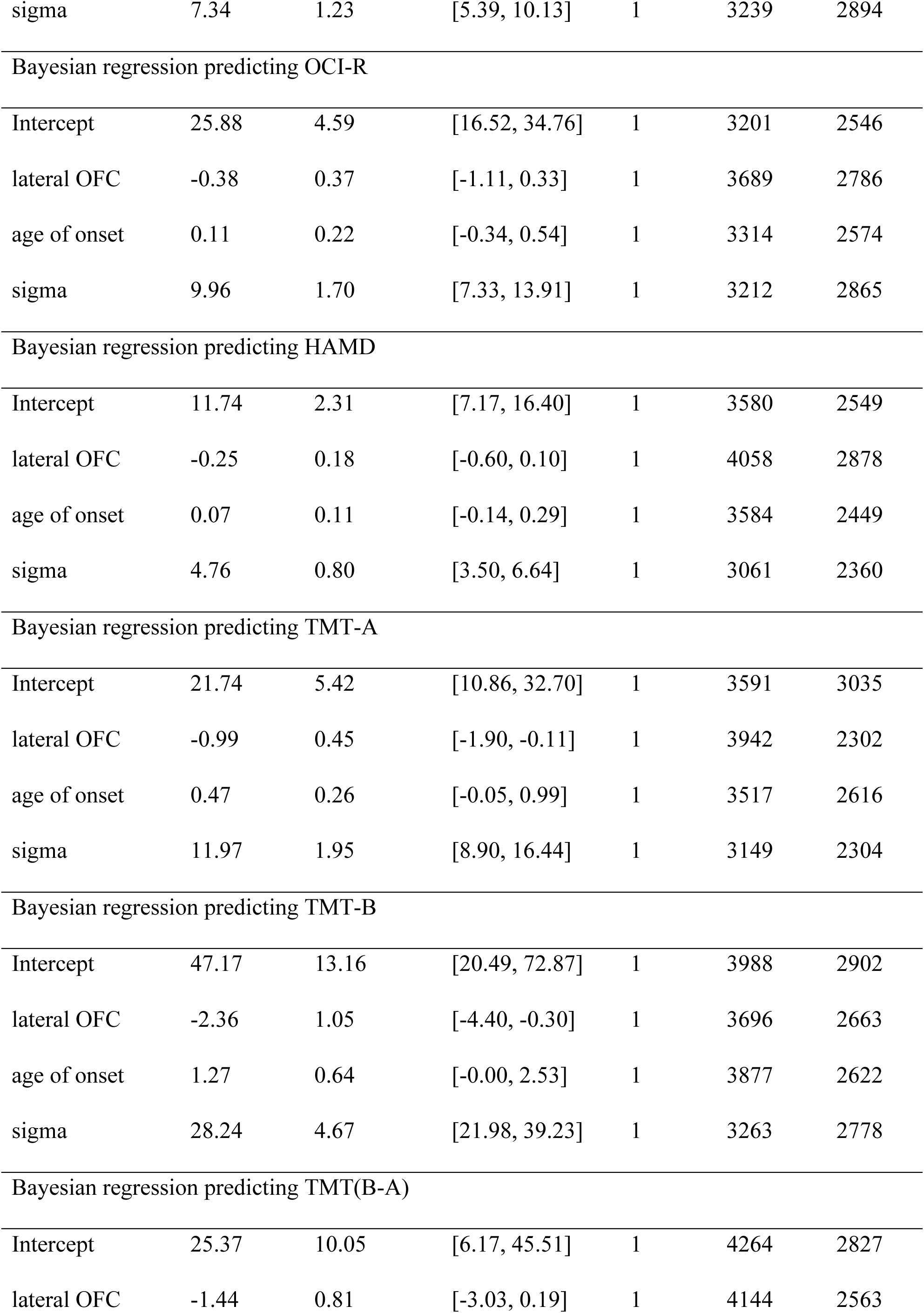
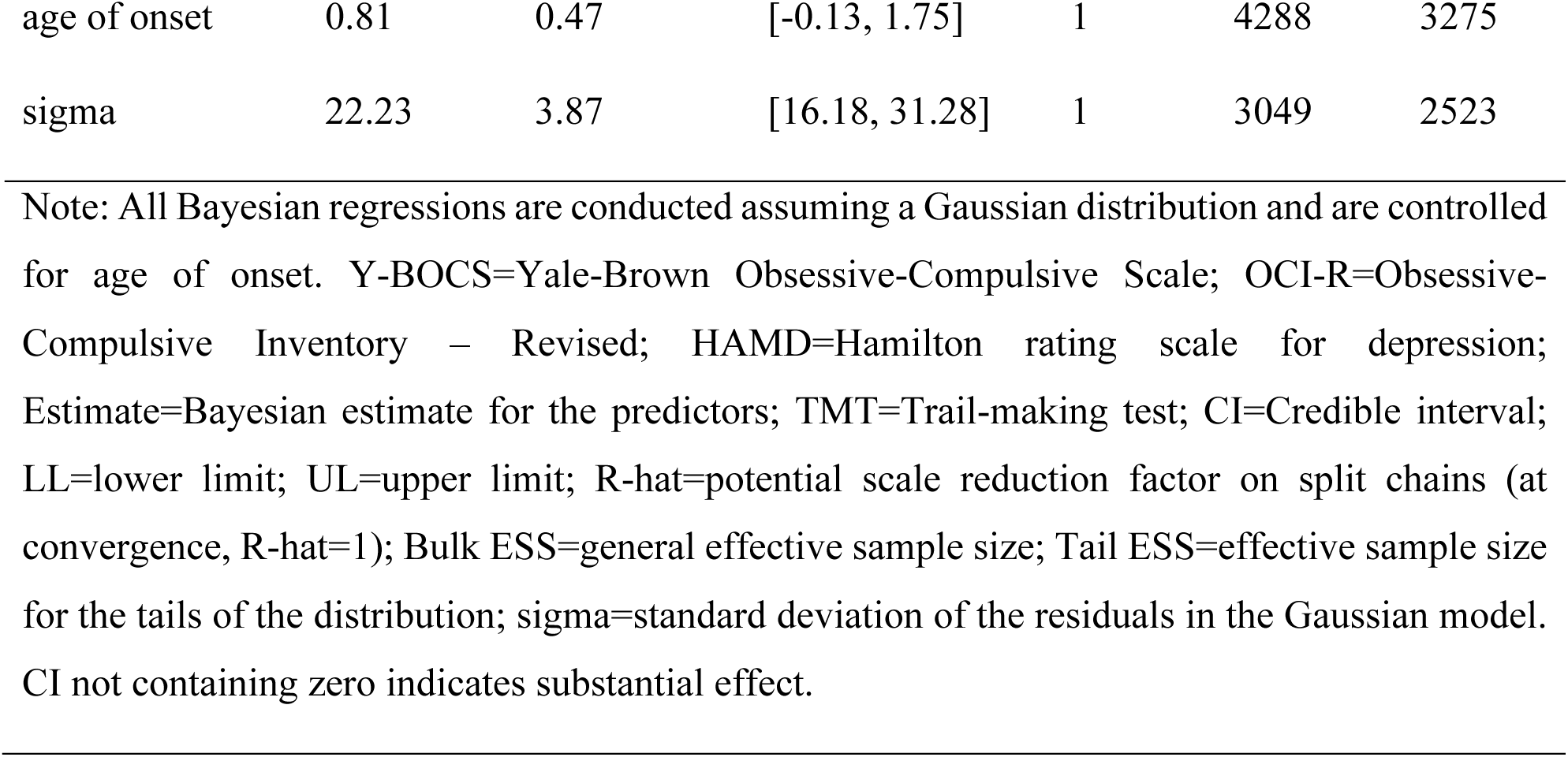
Model-based activations in the mPFC in OCD predicting clinical and cognitive scores using Bayesian linear regression analysis.

Please refer to the supplementary material, Section 2.7. for Bayesian correlations within the symptom scores.

## 4. Discussion

We investigated neural correlates of model-free and model-based prediction errors in OCD patients versus healthy controls using an fMRI-version of the two-step Markov decision task and computational modelling. Both groups relied on model-free strategies. Behaviorally, the model parameters revealed that the patients were more random, task performance revealed that they received fewer rewards than healthy individuals, and the stay probability results revealed that they relied more strongly on model-free compared to model-based strategies, which confirms previous findings (Banca et al., 2015; Knolle, Sen, et al., 2024; Voon et al., 2015a). Please refer to supplementary material Section 2.6. for detailed explanation of stay tendencies observed in the study. Additionally, refer to Supplementary Figure 10 for a comparison of the variability in the perseverance parameter estimates and the observed behavioral stay probabilities. While the behavioral data, supporting past literature (Robbins, 2024; Robbins et al., 2024; Voon et al., 2015b), indicates a potential bias towards habitual behavior, our neural findings revealed increased lateral OFC activity during model-based decision-making in OCD patients compared to controls, but revealed no group differences for model-free RPE processing. This increased activity in the lateral OFC may indicate a potential alteration in the model-based system, and may represent a compensatory mechanism by which the lateral OFC attempts to offset deficits in habitual decision-making, potentially enabling patients to maintain some aspects of goal-directed behavior despite underlying impairments. Interestingly, this activity was also associated with lower obsessive symptoms and better cognitive functioning.

While healthy controls exhibited activations in the lateral OFC during both model-based and model-free decision-making, OCD patients did not show activation in this region during model-free decisions but demonstrated hyperactivation compared to healthy controls during model-based decisions. This finding aligns with previous literature, which consistently reports lateral OFC hyperactivation in OCD during decision-making (Kim, 2021; Stern et al., 2013; Ursu and Carter, 2009). For instance, Ursu and Carter (2009) observed hyperactivity in the lateral OFC of OCD patients in response to cues associated with high-conflict probes, reflecting ruminative preoccupation with potential negative outcomes. They also found this hyperactivity to be correlated with the severity of anxiety symptoms. Similarly, using a sequential two-choice Markov decision task, Kim (2021) demonstrated that lateral OFC and lateral PFC hyperactivity in OCD patients contributed to excessively stable arbitration biased toward the model-free system, driving imbalanced decision-making. The hyperactivity in the lateral OFC observed during model-based decisions in OCD patients, therefore, might reflect a compensatory mechanism aimed at managing heightened conflict or uncertainty in goal-directed processing, potentially contributing to the imbalance between goal-directed and habitual systems. Interestingly, hyperactivity of the lateral OFC has also been associated with compulsive psychopathology in major depressive disorder, with activation during major depressive episodes suggested to act as a compensatory mechanism for mitigating negative emotional responses (Fettes et al., 2017).

Consistent with these findings, the literature highlights that the lateral OFC plays a critical role in maintaining a representation of task structure, assigning credit during value-guided learning, and resolving conflicts between goal-directed and habitual decisions (Nogueira et al., 2017; Watson et al., 2018). The observed lack of lateral OFC activation during model-free decisions, paired with its increased activation during model-based decisions in OCD patients, further supports the notion that hyperactivity in this region might reflect impaired conflict resolution within the goal-directed system, inadvertently reinforcing habitual behaviors. This observation aligns with the broader hypothesis that an impaired goal-directed system underpins the habitual biases observed in OCD (Gillan, 2021; Nestadt et al., 2016; Robbins, 2022). However, future research should investigate this relationship further. Together, these results underscore the role of lateral OFC hyperactivity as a potential neural correlate of disrupted goal-directed behavior in OCD.

Moreover, lateral OFC activity for model-based RPE signaling in OCD patients was negatively correlated with obsessive symptoms and cognitive functioning. This association, wherein increased lateral OFC activity corresponds to reduced obsessions and improved cognitive function, suggests that the observed hyperactivity in this region may serve a protective role in OCD. While this hyperactivation may help mitigate deficits in habitual decision-making, it might not be sufficiently effective in improving task performance, as evidenced by OCD patients receiving fewer rewards compared to healthy controls. This could indicate that while the compensatory mechanism is active, it may be inadequate in overcoming the behavioral impairments typically observed in OCD. Ursu and Carter, (2009) reported that the lateral OFC hyperactivation in OCD correlated with severity of anxiety. Given that OCD individuals engage in repetitive behaviors to avoid feelings of anxiety stemming from obsessions (Geramita et al., 2020), it is plausible the lateral OFC hyperactivity creates a bias toward the model-free system reinforcing repetitive behaviors, which in turn might function as a maladaptive strategy to alleviate the distress caused by obsessive thoughts, thereby contributing to reduced obsessions.

Furthermore, tasks assessing cognitive flexibility have shown hypoactivation in the lateral OFC activity in OCD patients (Brem et al., 2012; Chamberlain et al., 2008; Rotge et al., 2010), which was linked to weaker task performance and slower response times (Brem et al., 2012; Remijnse et al., 2009). This also raises the possibility that increased lateral OFC activation observed in our study reflects an effort to maintain cognitive functioning. Since this hyperactivation appears to support better cognitive performance in OCD patients, it may enable them to achieve a relatively comparable model-based performance to that of healthy controls despite underlying cognitive impairments, potentially explaining the lack of observed behavioral differences. However, future studies are needed to explore these associations.

In terms of model-free decision-making, OCD patients exhibited similar activations to controls, except in the lateral OFC and left caudate, where we observed weak evidence for activation in the controls but no evidence for activation in these regions in the OCD patients. This pattern aligns with previous studies linking lateral OFC activations more strongly to goal-directed than habitual decisions (Gremel and Costa, 2013; Rolls et al., 2020). Similarly, Guida and colleagues (2022) reported fewer and less strong activations in the left caudate for habitual activities in everyday-life, corroborating our observations.

The overlapping regions in the controls and patients that reflected model-free RPE included activations in the bilateral nucleus accumbens and putamen, the ACC, the mPFC, the medial OFC, and the right caudate, which is in accordance with our hypothesis and the literature (Banca et al., 2015; Burguière et al., 2015; Burton et al., 2015; Calzà et al., 2019; Frank and Claus, 2006; Gillan et al., 2015; Graybiel and Rauch, 2000; Harrison et al., 2009; Huang et al., 2020; Milad and Rauch, 2012). For model-based decisions, while both groups exhibited activations in the mPFC, lateral OFC, right caudate and the ACC, OCD patients recruited additional regions, including the medial OFC, left caudate, bilateral putamen and left accumbens. Despite both groups displaying similar decision-making behaviors in the task, the observed difference in brain activation during model-based decisions provides further support for OCD patients having an intact habitual but an altered goal-directed system.

Importantly, OCD patients exhibited overlapping activations during both decision-making processes, which might indicate an over-recruitment of regions associated with habitual decision-making when they struggle to recognize the benefits of goal-directed strategies. Supporting this, a task evaluating habitual versus goal-directed control revealed that OCD patients had impaired awareness of their actions’ outcomes and were more susceptible to action slips (Gillan et al., 2011b). Furthermore, the right nucleus accumbens was activated exclusively during model-free RPE, suggesting that activity in this region could serve as a potential neural correlate for model-free decision-making in OCD.

### 4.1 Limitations

This study has several limitations. First, our moderate sample size did not allow to explore subgroups of patients with specific symptoms. Subsequent studies should aim to replicate the results in a larger sample allowing for a differentiation in habitual and goal-directed decision-making between several symptom subtypes (e.g., washers versus checkers). Second, contrary to previous findings (Daw et al., 2011; Economides et al., 2015; Voon et al., 2015a), both the groups relied on model-free decisions, and we were unable to elicit model-based decision-making in healthy controls, potentially due to a combination of task instruction, reward presentation and task duration (Akam et al., 2015; Feher da Silva and Hare, 2020). Moreover, it is possible that the task design, which may not have been sufficiently sensitive to encourage model-based strategies, limited our ability to detect the full behavioral implications of this compensatory hyperactivation. Future studies should replicate our results with more trials, potentially in a longitudinal framework to provide a more thorough understanding. Third, our analysis included both medicated and unmedicated patients. Future studies should aim for larger sample sizes to verify results in separate analyses for medicated and unmedicated individuals.

## 5. Conclusion

This study provides new insights into the neural mechanisms underlying decision-making in OCD, revealing hyperactivity in the lateral OFC during model-based prediction error processing. These findings suggest that this hyperactivation may reflect an altered goal-directed system, potentially compensating for heightened conflict or uncertainty, while also reinforcing habitual behaviors. Importantly, activation in this region was also associated with reduced obsessive symptoms and improved cognitive functioning, potentially serving a protective or compensatory role, which may enable patients to achieve a comparable behavioral performance to healthy controls despite underlying impairments. Taken together, lateral OFC emerges as a critical region in the imbalance between habitual and goal-directed decision-making in OCD, offering a potential target for future interventions. Non-invasive techniques, such as transcranial direct current stimulation in this region may be useful for optimizing treatment by enhancing cognitive flexibility and mitigating obsessive symptoms in OCD patients.

## Acknowledgements

We would like to thank the patients and their families as well as the healthy controls for their time and effort. Furthermore, we would like to thank Dr. Felix Brandl and Dr. Christian Sorg for their insights and support during data acquisition.

## Data availability

Data and code available upon reasonable request to the corresponding author.

