## Supplementary material for "Altered model-based prediction error signaling in the lateral orbitofrontal cortex in patients with obsessive-compulsive disorder: An fMRI study": suppl material

### 1. Behavioral Analysis

#### 1.1. Statistical Implementations

All statistical analysis was conducted using the R Statistical Software (version 4.1.1) (R Core Team, 2021), and imaging analysis was performed using Statistical Parametric Mapping software, SPM12 (Ashburner et al., 2021). Masks for region of interest analysis were created the using FMRIB Software Library (FSL) (version 6.0.5.2) (Jenkinson et al., 2012). Behavioral data was visualized using the ggplot2 package (version 3.4.2) (Wickham, 2016), the ggpubr package (version 0.6.0) (Kassambara, 2020), and the Hmisc package (version 4.5-0) (Jr et al., 2021). Shapiro-Wilks tests was conducted with the rstatix package (version 0.7.2) (Singmann et al., 2021). Bayesian linear regression and ANOVA analysis was carried out using the brms package (version 2.20.4) (Bürkner, 2017). Bayes factor analysis was conducted using the bayesFactor package (version 0.9.12-4.5) (Morey & Rouder, 2023). Bayesian Spearman's correlations were conducted using the correlation package (version 0.8.4) (Makowski et al., 2022). ROI analysis using BML technique was conducted using the RBA package in AFNI (version 1.1.4) (Chen et al., 2019).

Before assessing for group-comparisons, data was inspected for normality of distribution using Shapiro-Wilks test. Since none of the datasets met assumptions for normality (Shapiro-Wilks $p>0.05$), Kruskal-Wallis tests and Bayesian robust ANOVAs were conducted. Bayesian Spearman Rank tests for used for correlation analyses.

#### 1.2. Task Comprehension and Engagement

The task was conducted in three blocks and the participants conducted 25 practice trials which were not included in the main analysis. The participants were also explicitly instructed about the task structure and that there were preferential transitions between stages, although the exact transition probabilities were not disclosed. Moreover, after the 25 practice trials before the start of the task, the researcher in charge questioned the participant about the structure, which further confirmed that the participants had good understanding of the task.

To illustrate that participants remained engaged with the task throughout, we plotted cumulative reward accumulation across trials for each group (Supplementary Figure 1). While cumulative reward is not a direct measure of learning in the two-step task, since rewards are determined by independent Gaussian random walks, it serves as a general indicator that participants were interacting with the task meaningfully.

Supplementary Figure 1 shows a steady increase in cumulative reward across trials in both groups, including in the later blocks of the task, suggesting sustained task engagement.

**Supplementary Figure 1**. Reward accumulation across trials.

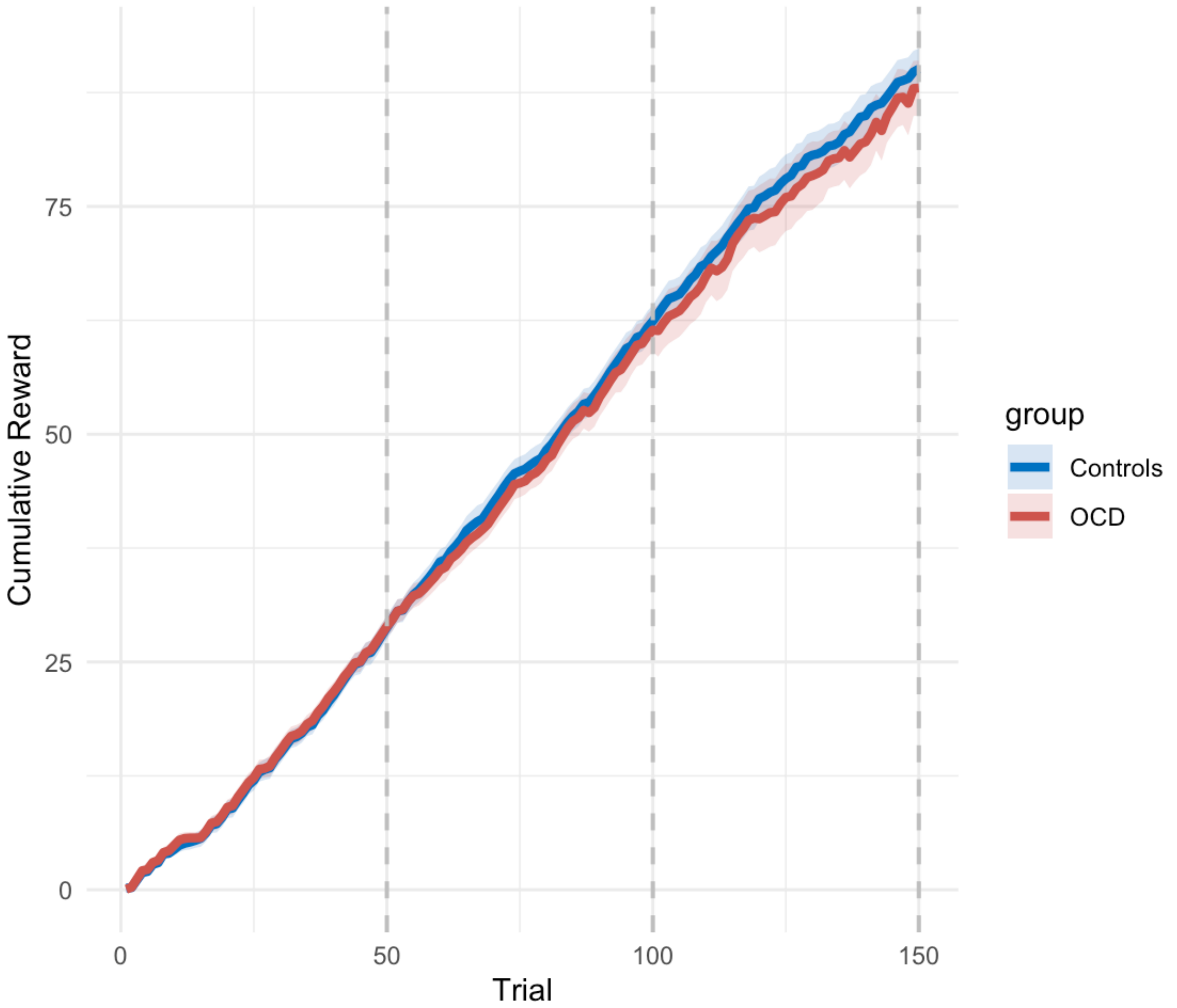


Note: The bold lines represent the mean cumulative reward for all participants in each group and the shaded regions represent 95% confidence intervals. The dashed vertical lines on trials 50, 100 and 150 represents the end of each block.

#### 1.3. Stay Probability

We examined the stay-probability on the first stage selection of each trial to see if the groups differed in their tendency for an action bias. Stay probability is the likelihood of an individual selecting the same initial stage choice as in the previous trial, and it is deterministic of whether the participant chooses a model-free or a model-based strategy for making decisions in the task.

Starting with the second trial, stay-probabilities were coded with 1 for stay and 0 for shift. If the previous trial resulted in a reward, model-free behavior would be characterized by repeating (stay: 1) the same choice on stage one on the following trial, and changing (shift: 0) the choice on stage one on the next trial if the previous trial did not result in a reward. Transition probabilities are not considered. Model-based behavior, on the other hand, would be characterized by repeating (stay: 1) the same choice on stage one at the next trial if the previous trial was either a common transition with a reward or an uncommon transition with no reward, and by changing (shift: 0) the choice on stage one at the next trial if the previous trial was either a common transition with no reward or an uncommon transition with a reward.

After assigning values to each trial, we estimated the percentage of '1's for model-based and model-free behavior four groups, namely, 'rewarded-common', 'rewarded-uncommon', 'unrewarded-common', and 'unrewarded-uncommon' for all participants and plotted histograms.

Stay probabilities were simulated for ideal model-based and model-free performances in order to visualize what the stay probabilities would be if all participants behaved totally model-based or model-free, and bar-plots were created. These plots were then visually compared with bar-plots made using stay probabilities computed according to the actual performance in the task.

#### 2. Computational Modelling

#### 2.1. Model specifications and description of models

We computationally modelled subjects' behavior across all subjects and also separately for patients and controls using the hBayesDM package (version 1.2.1) (Ahn et al., 2017) for R that uses Stan (version 2.26.3) to implement hierarchical Bayesian estimation. We fitted four chains for each model with 3000 iterations and 1000 burn-in samples. For further details please refer to Ahn and colleagues (2017). The models were fitted based on per trial information about stage 1 choice (1,2), stage 2 choice (1:4), and reward (0,1) as inputs. Model selection for further analysis was determined based on model-convergence and lowest leave one out information criterion (LOOIC) values. Descriptions of parameters generated by the models are presented in Supplementary Table 1.

**Supplementary Table 1.** Model descriptions

| Model | Description of parameters | Parameters |
|---|---|---|
| 4-parameter | As implemented by Wunderlich and colleagues (2012):<br>**Learning rate (‘a’):** efficiency of learning over the trials; higher the score, better the subjects understand and perform in the task<br>**Inverse-temperature/choice randomness (‘beta’):** influence of reward prediction on choices, beta referring to the proportions of random choices made during the task; beta = 0 : completely random responding and beta = ∞ : deterministically choosing the highest value option<br>**Perseverance (‘pi’):** how strongly the subject(s) stick to their decisions; higher the score, lower the chance of switching to a different image.<br>**Model-weights (‘w’):** degree of model-based influence on choices; w = 0 : more model-free decisions, and w = 1 : more model-based decisions. | a, beta, pi, w |
| 5-parameter | As implemented by Culbreth and colleagues (2016), in addition to **‘a’** and **‘pi’**, this model had **‘beta1MF’** and **‘beta1MB’** (choice randomness in stage 1 for model-free and model-based choices respectively), and **‘beta2’** (choice randomness for decisions in stage 2) | a, beta1MF, beta1MB, beta2, pi |
| 6-parameter | In addition to **‘beta1’** (choice randomness in stage 1, not separate for model-free and model-based choices), **‘beta2’**, **‘pi’**, and **‘w’**, this model included **‘a1’** and **‘a2’** referring to the learning rate of stage 1 and stage 2 respectively | a1, a2, beta1, beta2, pi, w |
| 7-parameter | An extension of the 6-parameter model as implemented in the study by Daw and colleagues (2011), included the above-mentioned six parameters along with **‘lambda’** (eligibility parameter), governing the relative importance of model-free and model-based reinforcer; λ = 1 where only the final reward is important, and λ = 0 where only the second-stage value plays a role | a1, a2, beta1, beta2, pi, w, lambda |

Note. Table showing parameter description, model fit and convergence for all the four models.

#### 2.2. Model architecture

Model-free value is denoted as $V_{s1}^{MF}$ and the model-based value $V_{s1}^{MB}$ for first stage stimuli s1 ∈ [1,2]. The model computes the actual value that is used in determining choice as weighted linear combination

$$V_{s1}^{HYBRID} = w \cdot V_{s1}^{MB} + (1 - w) \cdot V_{s1}^{MF}$$

Values for the four stimuli at the second stage (stimuli s2 ∈ [3..6]) are updated identically for both models according to reward prediction errors (Rummery & Niranjan, 1994):

$$V_{s2(t+1)} = V_{s2(t)} + \alpha_2\left(r - V_{s2(t)}\right)$$

At the first stage, model-free 'cached' values are updated according to temporal difference learning with reward prediction errors and eligibility traces:

$$V_{s1(t+1)}^{MF} = V_{s1(t)}^{MF} + \alpha_1\left(V_{s2chosen(t)} - V_{s1(t)}^{MF}\right) + \lambda \cdot \alpha_1\left(r - V_{s1(t)}^{MF}\right)$$

where $\alpha_1/\alpha_2$ are learning rates at the first and second stage, and $\lambda$ is a gain parameter for the eligibility traces.

Model-based values are calculated anew for each and every trial in a forward looking manner by multiplying the state values of the better option at the second stage with the state transition probabilities:

$$V_1{}^{MB} = 0.7 \cdot \max(V_3, V_4) + 0.3 \cdot \max(V_5, V_6)$$

$$V_2{}^{MB} = 0.3 \cdot max(V_3, V_4) + 0.7 \cdot max(V_5, V_6)$$

The probability P of choosing stimulus 1 (in a choice between stimulus 1 with value $V_1$ and stimulus 2 with value $V_2$) is computed in stage 1 according to a softmax choice function dependent on the relative stimulus values and choice $C$ in the previous trial.

$$P(1) = \frac{1}{\{1 + exp( -\beta_1(V_1 - V_2) - \pi(C_1 - C_2))\}}$$

and similarly, in stage 2

$$P(1) = \frac{1}{\{1 + exp( -\beta_2(V_1 - V_2))\}}$$

For the 5-parameter model to predict first-stage choices, model-based and model-free action values were combined to calculate action probabilities using a softmax function:

$$P(1) = \frac{1}{\{1 + \exp( -\beta_1^{MF}(V_1 - V_2) - \beta_1^{MB}(V_1 - V_2) - \pi(C_1 - C_2))\}}$$

For additional in-depth information on the computational model, please refer to Daw et al., (2011), and Brandl et al., (2023).

#### 2.3. Model selection

We first fitted the four models across all subjects a single hierarchical prior across all subjects, independent of group to allow for direct comparison across the groups. Using a single prior reduces the risk of overfitting, particularly given the relatively small sample sizes, and improves the stability of parameter estimates.

Moreover, such a conservative approach requires the group differences to be consistent across participants for effects to emerge, meaning that any effects detected in the analysis are most likely robust. By using this approach, we also avoid biasing our results to identify group differences by fitting separate priors for each group.

While group-specific priors might better capture subtle differences, such an approach would require larger sample sizes to ensure model convergence. Nevertheless, to explore potential differences, we also ran all four Bayesian models separately for the groups. However, these models did not converge well, likely due to the small sample sizes within each group. Among the models, Model 6, which was fitted across all participants, was found to be the best-fitting model based on both model convergence and LOOIC values, and was chosen for further computational analysis.

A summary of the model fits (LOOIC values) and the caterpillar plots showing model convergence for all the models run separately for each group, and across all participants, are presented below in Supplementary Table 2 and Supplementary Figures 2-4. If the caterpillar plots are not skewed at one end, indicating no evident divergence or drift; and if straight line can be drawn through the middle without the line overlapping any other chain, indicating that the chains have mixed, it can be stated that the model has converged. This, along with the lowest LOOIC values determined the best fitting model (6-parameter model run across all subjects).

**Supplementary Table 2 .** LOOIC table

| Model | LOOIC | | |
|---|---|---|---|
| | OCD patients | Healthy Controls | Across all |
| Model 4 | 7459.27 | 5942.833 | 13373.14 |
| Model 5 | 7386.2 | 5855.236 | 13222.703 |
| Model 6 | 7289.762 | 5782.502 | 11744.329 |
| Model 7 | 7287 | 5793.794 | 13051.582 |

Note. Table showing model fit for all the four models. LOOIC = Leave one out information criterion.

**Supplementary Figure 2**. Model convergence diagnostics for 4-, 5-, 6-, and 7-parameter models fit to healthy control data.

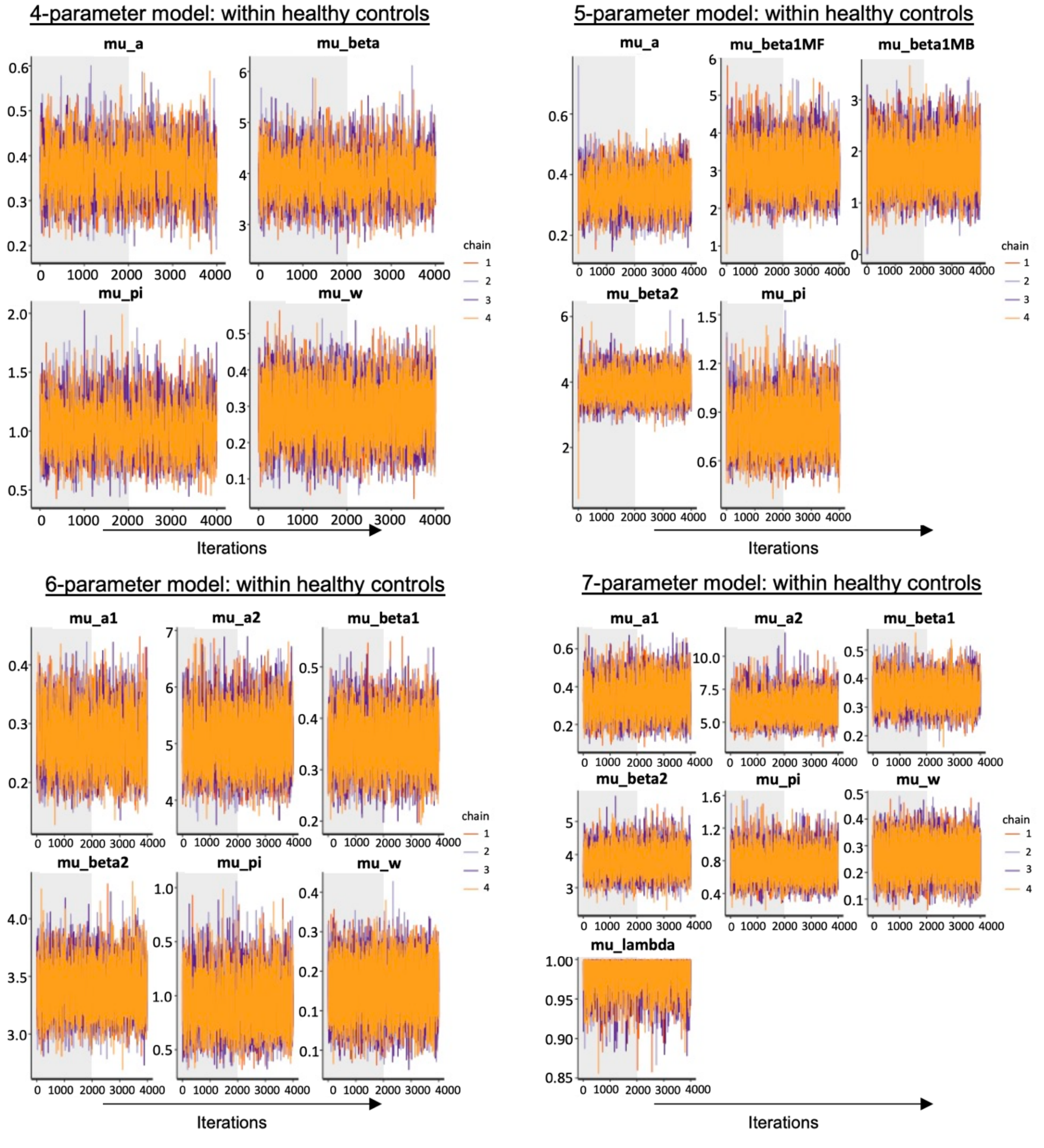


Note: Trace plots showing MCMC sampling of group-level (hyper) mean parameters for hierarchical Bayesian models fit to data from healthy controls. Each panel represents a parameter trace across iterations for four sampling chains (color-coded). Shaded grey areas indicate the burn-in period. The models vary in complexity, including 4-, 5-, 6-, and 7-parameter specifications. Parameter names for each model follow the conventions provided in Supplementary Table 1, where the prefix "mu_" indicates the group-level (hyper) mean of each individual-level parameter (e.g., "mu_a" corresponds to the group-level mean of individual learning rate parameter a).

**Supplementary Figure 3.** Model convergence diagnostics for 4-, 5-, 6-, and 7-parameter models fit to OCD patient data.

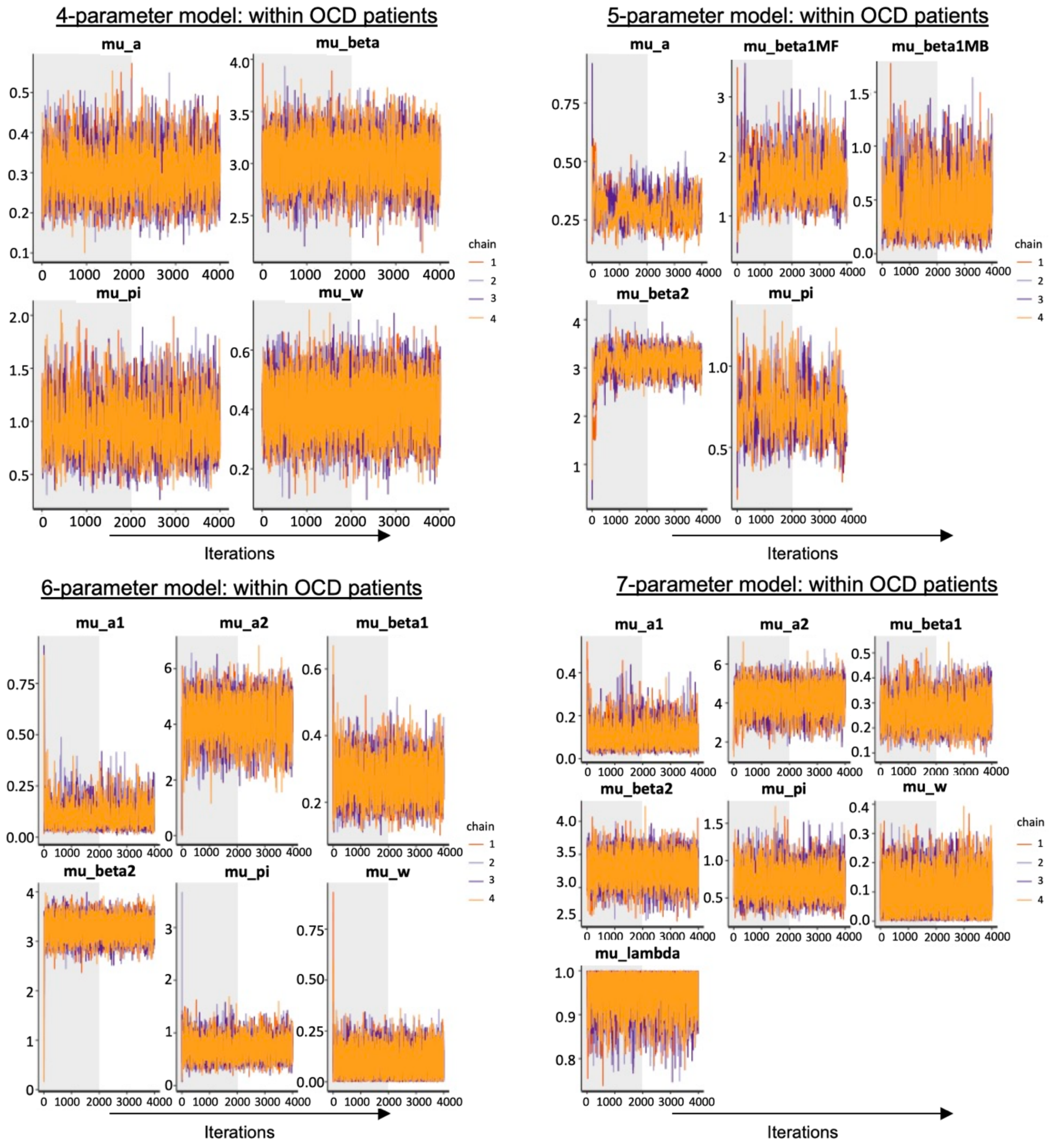


Note: Trace plots showing MCMC sampling of group-level (hyper) mean parameters for hierarchical Bayesian models fit to data from OCD patients. Each panel represents a parameter trace across iterations for four sampling chains (color-coded). Shaded grey areas indicate the burn-in period. The models vary in complexity, including 4-, 5-, 6-, and 7-parameter specifications. Parameter names for each model follow the conventions provided in Supplementary Table 1, where the prefix "mu_" indicates the group-level (hyper) mean of each individual-level parameter (e.g., "mu_a" corresponds to the group-level mean of individual learning rate parameter a).

**Supplementary Figure 4.** Model convergence diagnostics for 4-, 5-, 6-, and 7-parameter models fit across all subjects.

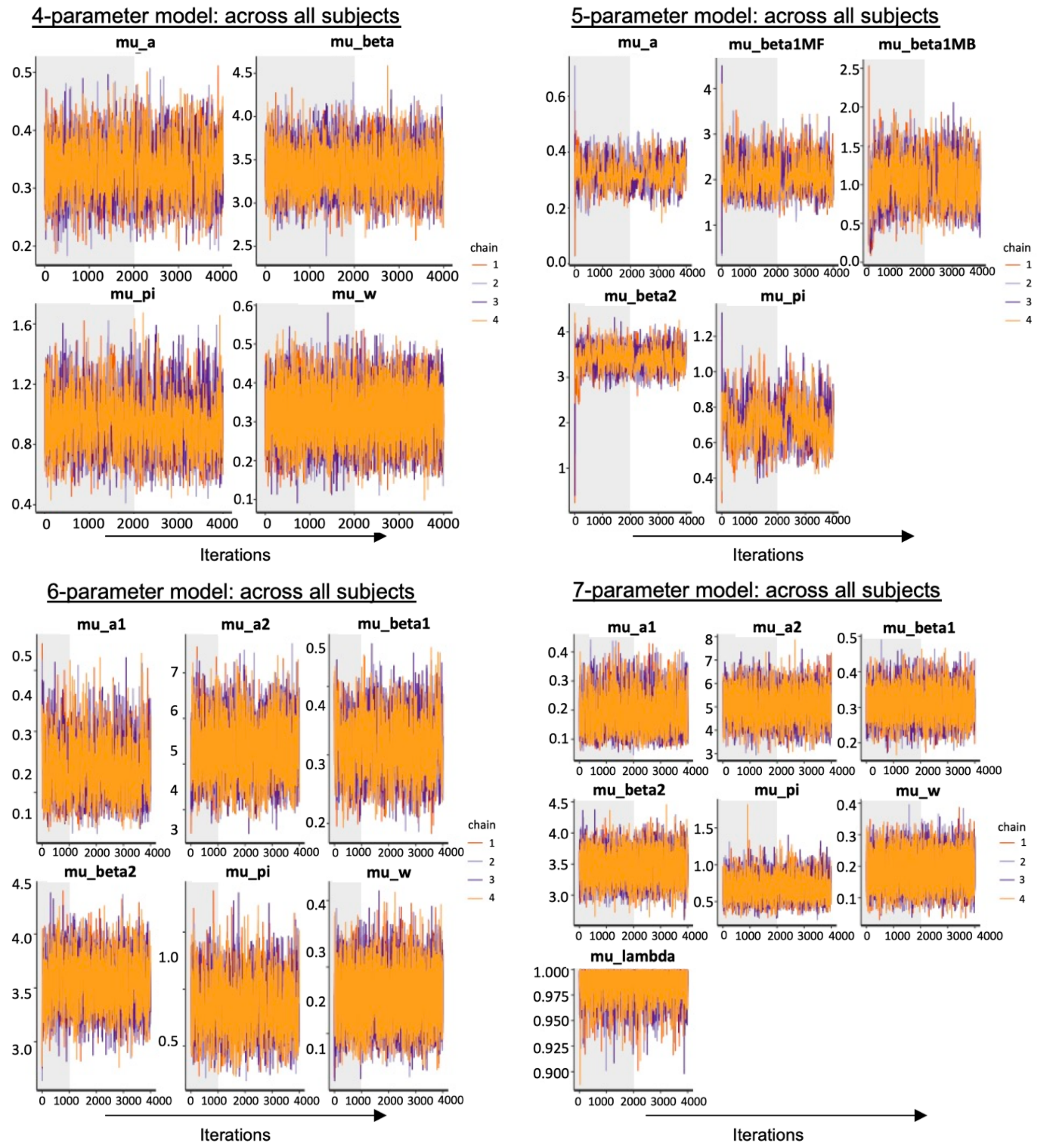


Note: Trace plots showing MCMC sampling of group-level (hyper) mean parameters for hierarchical Bayesian models fit to data from all the subjects. Each panel represents a parameter trace across iterations for four sampling chains (color-coded). Shaded grey areas indicate the burn-in period. The models vary in complexity, including 4-, 5-, 6-, and 7-parameter specifications. Parameter names for each model follow the conventions provided in Supplementary Table 1, where the prefix "mu_" indicates the group-level (hyper) mean of each individual-level parameter (e.g., "mu_a" corresponds to the group-level mean of individual learning rate parameter a).

Box plots showing the distribution of individual parameter estimates for the two groups from the winning model (Model 6 run across all subjects) are displayed below in Supplementary Figure 5.

**Supplementary Figure 5.** 6-parameter model parameters run across all subjects.

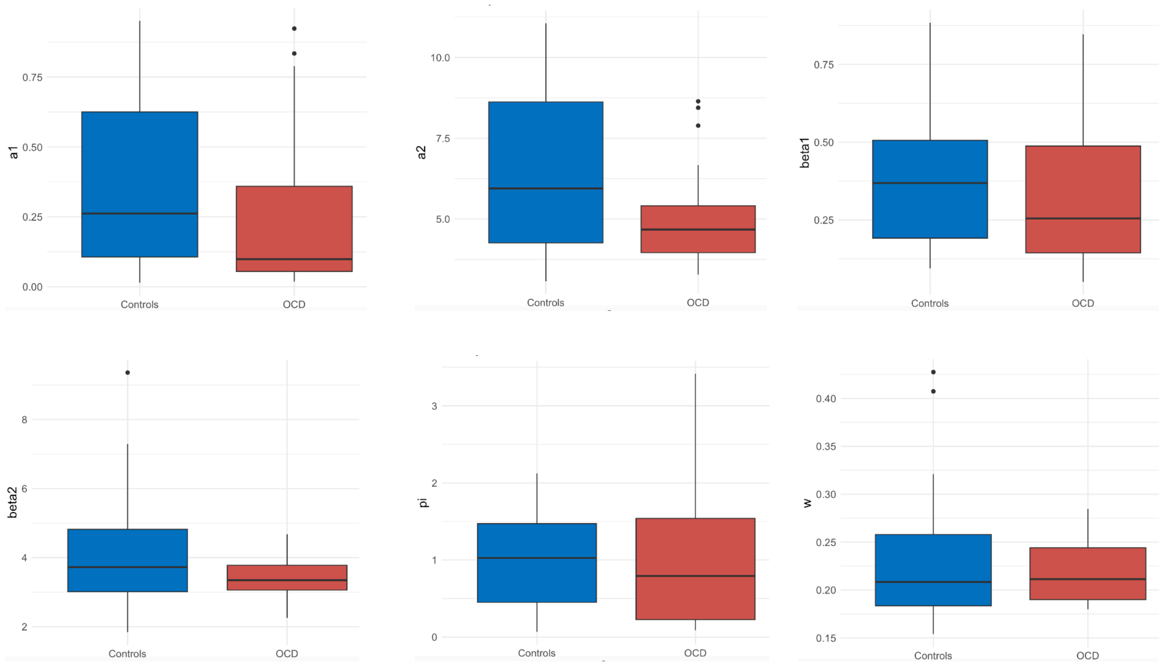


#### 2.4. Model Regressors

The hBayesDM package allows the users to extract trial-by-trial latent variables (e.g., prediction error) required for model-based fMRI/EEG analysis as parametric modulators. However, the two-step hBayesDM models did not have model regressors available to use as parametric modulators for imaging analysis, therefore, we imported the codes for the models from the publicly available github repository (https://github.com/ccs-lab/hBayesDM) and made changes to the .stan and .yaml files to include model-based and model-free regressors with the help of the following equations based on the study by Daw and colleagues (2011):

- mf_RPE[i, t] = reward[i, t] - v_mf[level1_choice[i, t]]; // regressor for model-free reward prediction error
- mfb_RPE[i, t] = (reward[i, t] - v_mf[level1_choice[i, t]]) - (reward[i, t] - v_mb[level2_choice[i, t]]); // regressor for difference between model-free and model-based reward prediction error

where, mf_RPE = model-free reward prediction error for each trial for each subject,

mfb_RPE = model-based reward prediction error for each trial for each subject,

reward = reward received at every trial for every participant,

v_mf = model-free stimulus values for level 1&2,

v_mb = model-based stimulus values for level 1,

i = subject ID,

t = trial number

The rationale behind using a difference regressor instead of the simple model-based regressor was to isolate the neural signal uniquely attributed to model-based processes while minimizing confounding contributions from model-free systems. Prediction error signals in the brain often reflect overlapping contributions from both model-free and model-based systems, making it challenging to interpret the respective contributions of each process. This overlap can be seen in the plots below for model-free and model-based regressors, which show strikingly similar patterns across trials (Supplementary Figures 6 and 7), further illustrating that a simple model-based regressor may not fully capture true model-based processes. Without accounting for this shared variance, the neural signals regressed on the model-based regressor might also reflect model-free influences. Therefore, to disentangle the contributions of the model-based system, we adopted a difference regressor that isolates the true model-based signal, as explained by Daw et al., (2011). In their work, the authors calculated a difference regressor by subtracting model-free prediction errors from the model-based RPEs. The two key reasons for this approach are:

1. Reducing regressor correlation: Including both model-free and model-based regressors independently in the analysis could lead to multicollinearity, as these regressors are often highly correlated as shown in the first two plots below. The difference regressor mitigates this issue by reducing the shared variance between the two signals.

2. Isolating unique neural contributions: As Daw et al., (2011) explain, this approach allows for testing whether the BOLD activity is purely model-free, purely model-based, or a combination of both. If the signal is entirely model-free, it will be captured by the model-free regressor, and the difference regressor will not explain additional variance. Conversely, if the signal reflects any model-based influence, the difference regressor

will account for this residual variance, indicating unique contributions from the model-based system.

We implemented a similar approach by calculating the model-based difference regressor as the difference between the model-free RPE and the model-based RPE. This regressor isolates the neural component uniquely attributable to model-based processes, removing contributions from the model-free system. The resulting time-course of the difference regressor exhibits a distinct pattern that diverges from both the model-free and model-based regressors, as shown in the plots below. This distinctiveness demonstrates that the difference regressor successfully isolates the unique signal associated with model-based processes (Supplementary Figure 8).

The initial values of the regressors in the plots below provide additional insight into the dynamics of model-free and model-based learning. The model-free regressor starts at a positive value (around 0.2) (Supplementary Figure 6), reflecting a prediction error signal generated by the model-free system based on its default expectations of reward at the beginning of the task. In contrast, the difference regressor starts at zero (Supplementary Figure 8), indicating the absence of a significant model-based contribution on the first trial. This suggests that early in the task, behavior is predominantly driven by model-free learning, as the model-based system requires experience to learn the transition structure and begin contributing to predictions. This interpretation aligns with established reinforcement learning frameworks where model-free processes dominate during initial learning stages, providing further support for the difference regressor reflecting true model-based behavior.

**Supplementary Figure 6.** Model-free regressor (mf_RPE) across trials

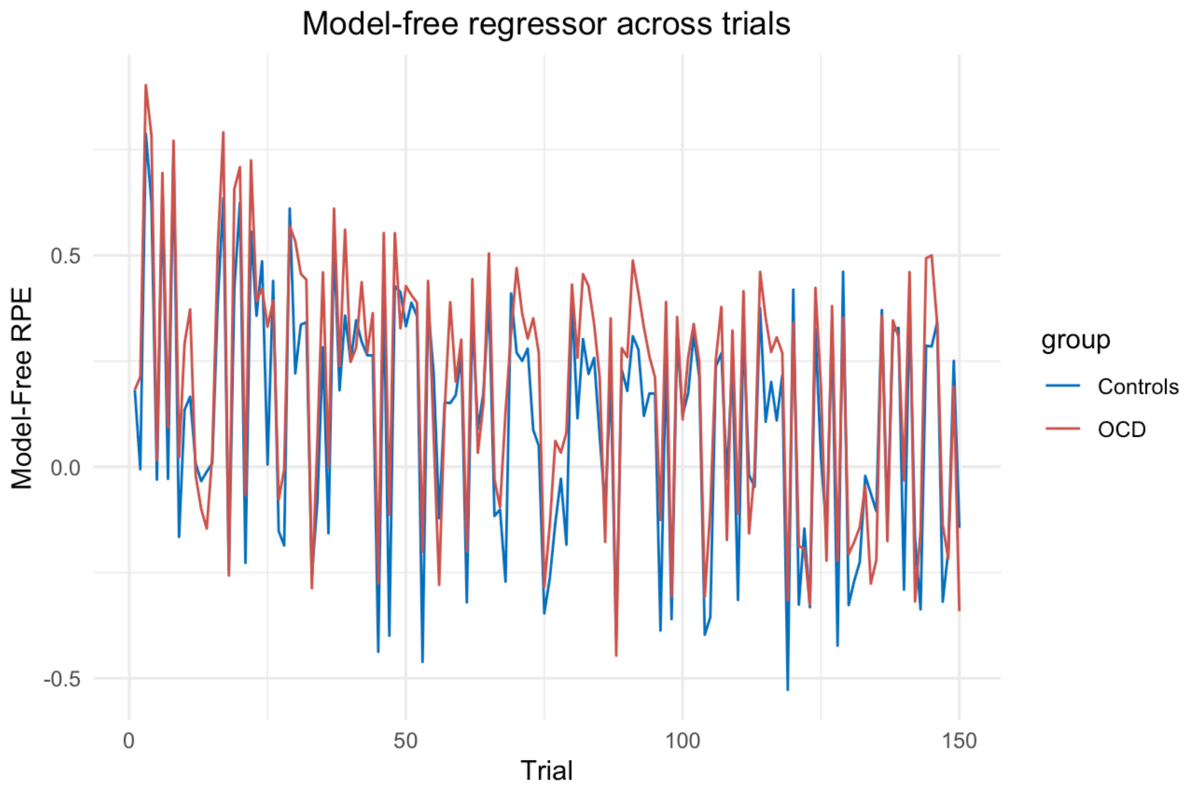


**Supplementary Figure 7.** Model-based regressor across trials

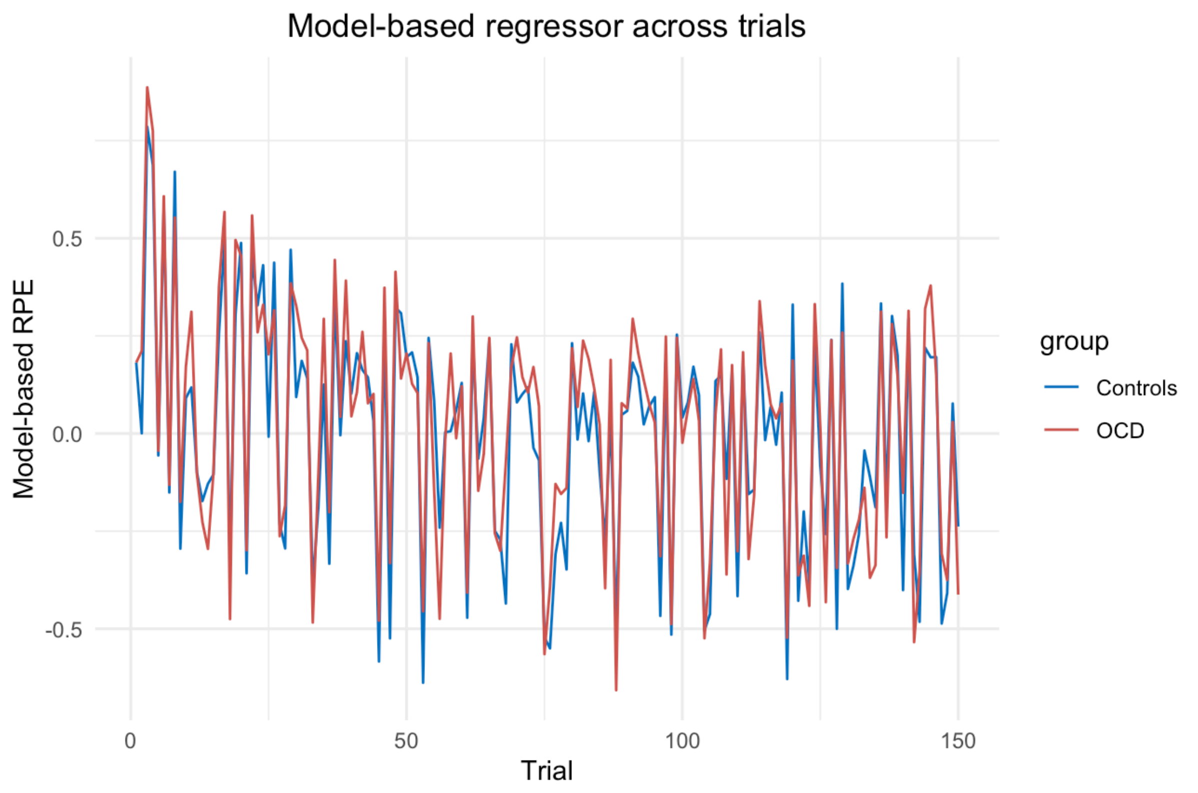

**Supplementary Figure 8.** Model-based difference regressor (mfb_RPE) across trials

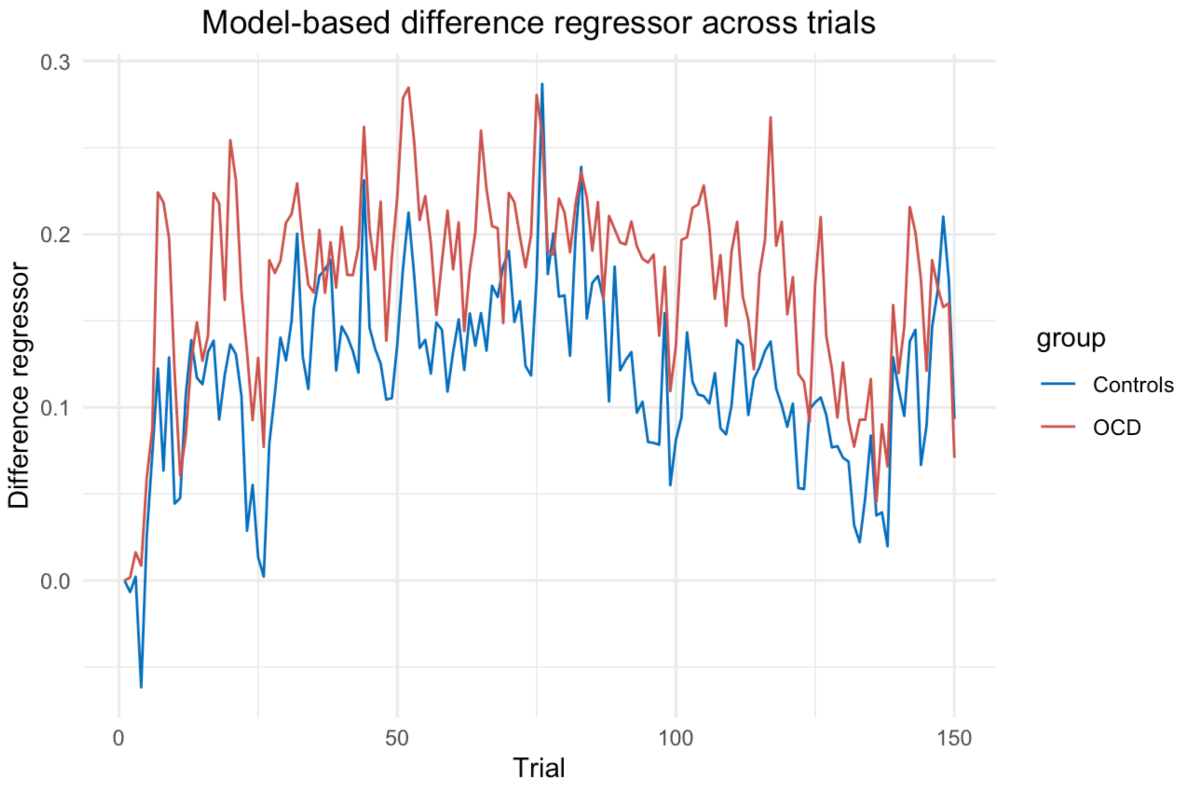


#### 2.5. Variance in behavior explained by computational model

To quantify how much variance in behavior the winning 6-parameter hierarchical Bayesian model explains, we performed a Bayesian regression analysis to see how well the model parameters explain the stay probability behavior. We also incorporated a subject level intercept to account for individual differences in behavior.

Model:

$$stay\_probability \sim \alpha_1 + \alpha_2 + \beta_1 + \beta_2 + \pi + w + (1 \mid SubjID)$$

where, $\alpha_1$ & $\alpha_2$ are learning rate stage 1 and 2 respectively, $\beta_1$ & $\beta_2$ are inverse temperature/choice randomness stage 1 and stage 2, $\pi$ is perseverance, and $w$ is model-weights.

This model explained 93.75% of the variance in the observed behavior (Bayesian $R^2 = 0.9375$, EE = 0.0393, 95% CI [0.8695, 0.9968]), suggesting that it effectively captures the relationship between the observed behavior and the model-derived parameters. The narrow credible interval and the small estimated error further supports the model's strong explanatory power.

This analysis revealed that the model accounts for a substantial proportion of variance in behavior, providing confidence in its explanatory power.

Additionally, the random intercept for participants (‘SubjID’) showed a moderate variability in baseline stay probability (B = 0.04, 95% CI [0.00, 0.07]), suggesting that individual differences in baseline behavior exist but are not large. The population-level effects indicated several relationships between the predictors and stay probability. Specifically, learning rate in stage 1 ($\alpha_1$: B = 0.16, EE = 0.03, 95% CI [0.10, 0.22]), learning rate in stage 2 ($\alpha_2$: B = 0.03, EE = 0.00, 95% CI [0.02, 0.04]), and perseverance ($\pi$: B = 0.17, EE = 0.01, 95% CI [0.15, 0.20]) positively predicted stay probabilities.

Following is the plot (Supplementary Figure 9) for the posterior predictive check of the above Bayesian regression analysis where y represents the actual stay probability values and $y_{rep}$ represents model-replicated stay probability values, further supporting the model’s validity.

**Supplementary Figure 9.** Posterior predictive check for Bayesian regression model predicting stay probability from computational model parameters.

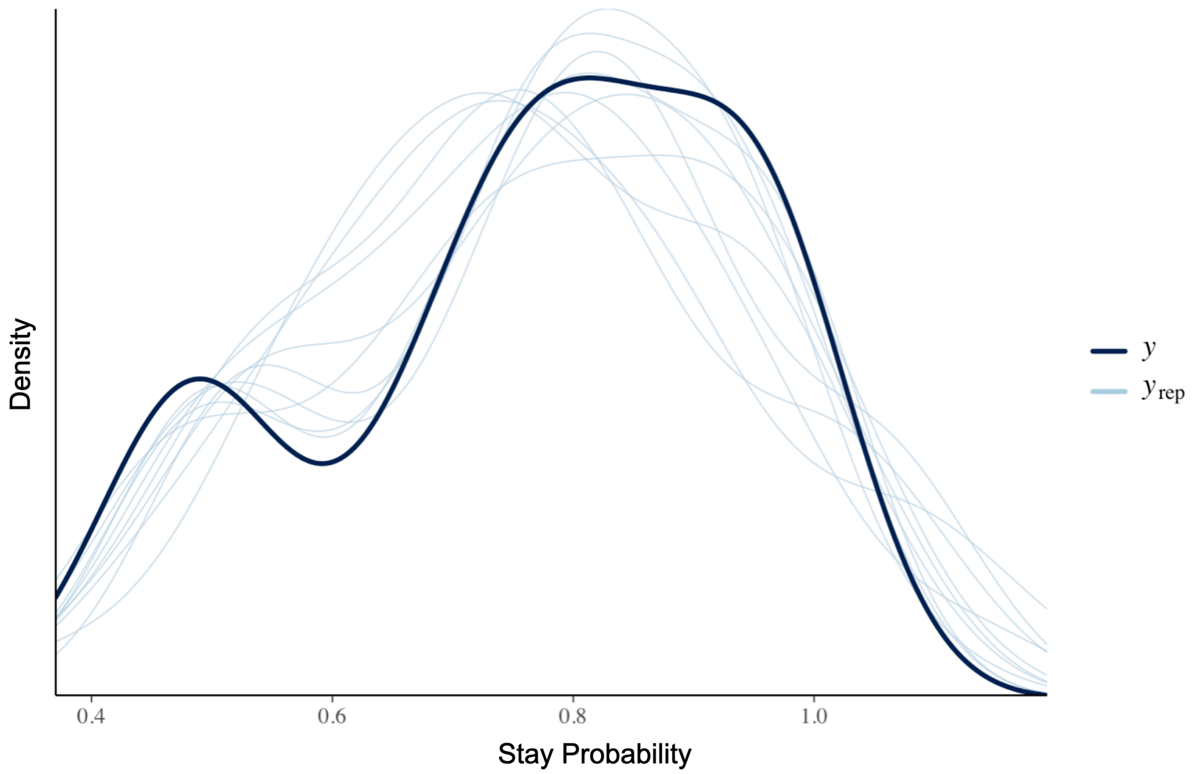


Note: The dark line (y) represents the observed distribution of stay probabilities, while the lighter lines ($y_{rep}$) represent model-replicated distributions generated from posterior samples. The close alignment between observed and replicated data indicates a good model fit.

These checks demonstrate that the model provides a good approximation of participants' behaviors and effectively capturing decision-making patterns across all subjects.

#### 2.6. Stay tendencies – Perseverance and Stay Probability

The perseverance parameter explains the tendency to stick to the same first level choice independent of reward. It captures the participants' tendency to repeat previous choices, irrespective of outcomes. In our study, we observed that the perseverance parameter values were relatively low in both groups, indicating a weak overall tendency to stay on the same first-level choice. Both groups show similar perseverance values (pi) with comparable averages as shown in the boxplot presented below (Supplementary Figure 10) showing the model parameter distributions for both groups.

The model parameter perseverance, however, provides a more sensitive measure of inter-individual variability compared to simple behavioral stay probabilities because it accounts for patterns of behavior across the entire task taking into account the task structure, rather than being limited to single-trial effects. Thus, while behavioral stay probabilities are calculated based on the response to the immediate prior trial, the perseverance parameter integrates information from cumulative performance, allowing for a more comprehensive and nuanced assessment of an individual’s tendency to repeat previous choices irrespective of reward.

Although in our behavioral stay probability analysis we found that the participants had a strong tendency to stay on the same first level choice if the previous trial was rewarded, the perseverance parameter values were relatively low in both groups, suggesting only a weak overall tendency to stay. Moreover, we did not observe any group differences in the perseverance parameter. This finding aligns with previous studies using the same task in control and OCD patients, which also reported low perseverance values, lack of significant group differences in perseverance, and similar behavioural stay probabilities like our study (Knolle, Sen, et al., 2024; Voon et al., 2015). These findings suggest that the overall tendency to stay in this task is uniformly low across both groups, indicating more flexible or adaptive decision-making style.

While the perseverance parameter provides a more comprehensive measure of participants’ tendency to stay compared to single-trial behavioral stay probabilities, it is a valid concern that the relatively low parameter values could obscure individual differences. To reliably explore such differences, larger sample sizes and multiple timepoints would be necessary to ensure replicability and robust variability.

Additionally, to assess the variability captured by the perseverance parameter compared to behavioral stay probabilities, we compared their respective variances as shown in the boxplot below in Supplementary Figure 10. The perseverance parameter demonstrates greater variability than behavioral stay probabilities across participants, supporting its potential utility for capturing inter-individual differences. Visual inspection of the parameter distributions also suggests slightly greater variability in perseverance values among OCD patients compared to controls, which may reflect greater heterogeneity within the OCD group. Future studies should aim to explore this potential variability further, and replicate these findings with larger and more diverse samples to ensure a more thorough evaluation of inter-individual differences.

**Supplementary Figure 10.** Variances in model parameter perseverance and behavioral stay probability

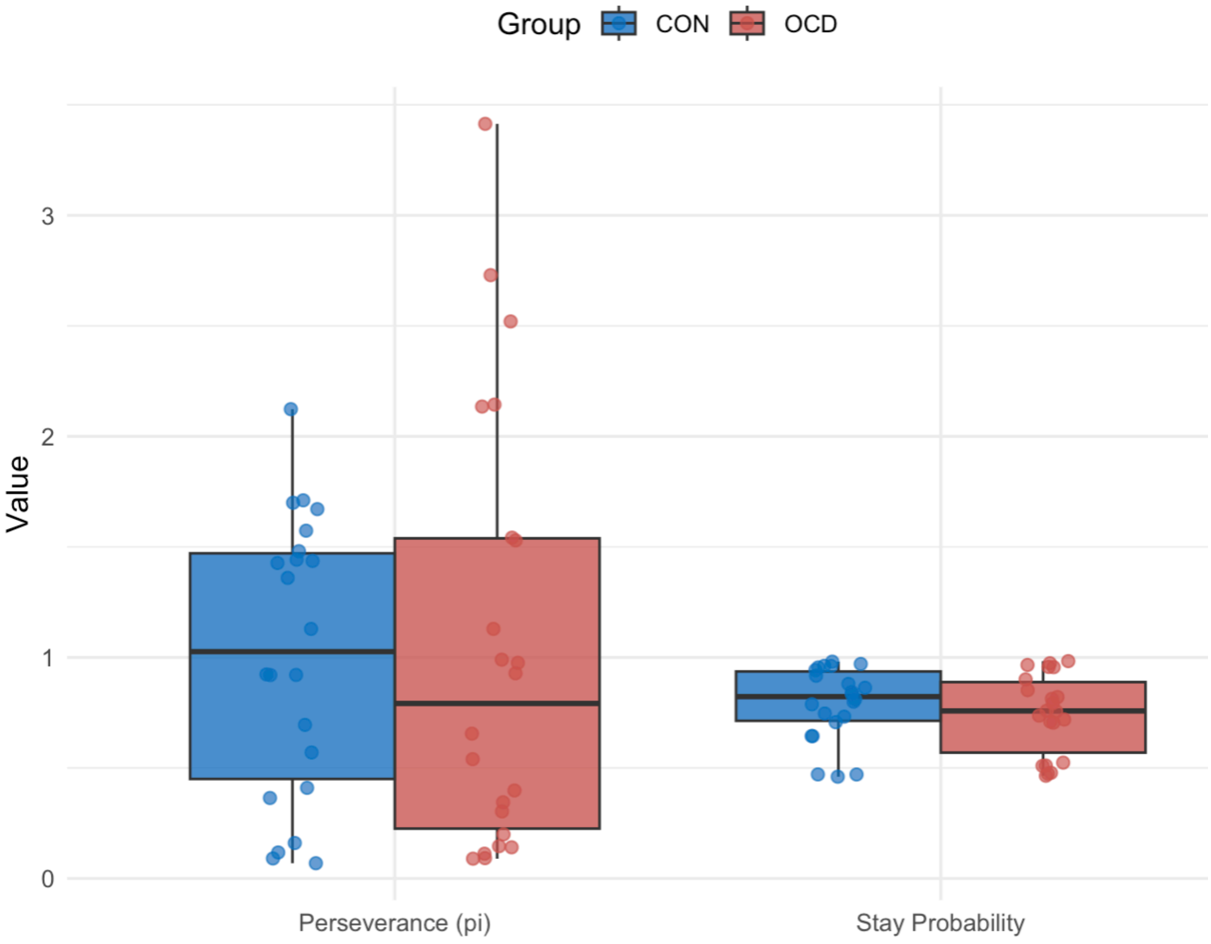


#### 2.7. Within symptom correlations in OCD patients

In line with past observations of compulsive behaviors to be linked to metacognitive impairments (Gillan, 2021), we also observed positive correlations between Y-BOCS compulsions and TMT-B (rho = 0.34, 95% CI [0.00, 0.67], BF10 = 2.13), and Y-BOCS total and TMT-B (rho = 0.38, 95% CI [0.01, 0.68], BF10 = 3.41), indicating that higher compulsive and overall OCD symptoms are linked to slower executive functioning and worse cognitive flexibility. Additionally, OCI-R was found to be positively correlated with both TMT-A (rho = 0.39, 95% CI [0.05, 0.69], BF10 = 3.74) and TMT-B (rho = 0.37, 95% CI [0.04, 0.69], BF10 = 3.56), indicating that overall OCD symptom severity is also linked to slower processing speed, executive functioning and worse cognitive flexibility.
